# Increased risk of asthma in female night shift workers

**DOI:** 10.1101/2024.12.05.24318539

**Authors:** Robert J. Maidstone, David W. Ray, Junxi Liu, Jack Bowden, Martin K. Rutter, Hannah J. Durrington

## Abstract

**Background:** Asthma is more common in females and more common in night shift workers. Since increasing numbers of females are becoming shift workers it is important to determine if shift work-associated asthma risk is higher in females.

**Research Question:** Is increasing frequency of night shift work more strongly related to prevalent asthma in females than in males?

**Study Design and Methods:** We used cross-sectional data from >280,000 UK Biobank participants and logistic regression models adjusted for demographic and lifestyle factors to describe sex differences in prevalent asthma related to shift work frequency. To obtain mechanistic insights, we explored associations with chronotype, sex hormones and menopause.

**Results:** Compared to female day workers, female permanent night shift workers had higher covariate-adjusted odds of moderate-severe asthma (OR: 1.50 (95% CI 1.18 – 1.91)) but there was no corresponding relationship in males (OR 0.95 (95% CI 0.72 – 1.26); sex interaction p-value = 0.01). Similar relationships were observed for ‘all asthma’ and for ‘wheeze or whistling in the chest’ outcomes. Female shift work-related asthma was driven by relationships in postmenopausal women not using HRT (e.g., adjusted OR: 1.89 (1.24-2.87) for moderate-severe asthma; sex interaction p-value = 0.02 in permanent nightshift workers compared to dayworkers) but these relationships attenuated to the null in postmenopausal women using HRT.

**Interpretation:** Our finding that increasing shift work frequency is more strongly related to asthma in females than in males could have Public Health implications. Intervention studies should determine if modifying shift work schedules or HRT can reduce asthma risk in females.

---

Asthma is a highly rhythmic inflammatory disease with symptoms worsening overnight (1), coinciding with exaggerated airway narrowing and increased airway inflammation (2).

Using data from more than 280,000 UK Biobank participants (3) we previously showed that, compared to day workers, ‘permanent’ night shift workers had higher likelihood of moderate-severe asthma, possibly due to disrupted circadian rhythms.

Asthma predominantly affects females, who have more severe disease, higher hospitalisation and death rates compared to males (4). The higher asthma prevalence in females starts at puberty and is associated with fluctuations in hormones during menstruation, pregnancy, and menopause suggesting a possible causal role for female sex hormones in asthma pathogenesis (5).

Oestrogens regulate the circadian molecular clock (6), and oestrogen receptor β is expressed in a circadian manner, particularly in lung epithelial cells, so coupling circadian phase to oestrogen action in the lung (7). Compared to males, females tend to have an earlier chronotype, likely related to a shorter intrinsic circadian period (8, 9, 10).

Since night shift work is currently undertaken by ∼20% of workers (11) and 2/3 of the recent increase in shift worker numbers is in females (12), it is important to understand if shift work is linked to a higher risk for asthma in females than in males.

Using data on UK Biobank participants, our main aim was to investigate sex differences in the association between shift work and asthma. To obtain mechanistic insights, we explored relationships of chronotype and sex hormones with asthma, and how asthma risk in female shift workers is related to menopausal status.

## Study Design and Methods

UK Biobank recruited 502,540 participants (5% of those invited) aged 40 to 69 years (13). Our analysis was restricted to participants in paid employment or who were self-employed at baseline (N=274,541) (14).

### Cases of asthma

Cases of asthma and moderate-severe asthma were defined as previously (15). Briefly, we identified 13,589 (5.3% of our study cohort) asthma cases, of which 4,549 (1.9%) had moderate-severe asthma. We excluded participants with doctor-diagnosed asthma who did not report taking asthma medication and participants reporting taking asthma medication who did not report having doctor-diagnosed asthma (N=19,287). For analysis of moderate-severe asthma we further excluded those not receiving medication for moderate-severe asthma ((15); N = 9,040).

We also analysed data from participants who had ‘experienced wheeze or whistling in the chest within the last year’ and participants with obstructive pulmonary function tests (<80% of the predicted percentage FEV1 (forced expiratory volume in 1 second)).

### Shift work and chronotype

We categorized current shift work status as either: a) ‘day worker’, b) ‘shift worker, but only rarely if ever nights’, c) ‘irregular shift work including nights’, and d) ‘permanent night shifts’ from responses to the UK Biobank baseline questionnaire (15). Similarly, self-reported chronotype was categorized as either: a) “definitely a ‘morning’ person”, b) “intermediate chronotype” or c) “definitely an ‘evening’ person” (15).

### Sex hormones

Testosterone, sex hormone binding globulin (SHBG) and oestradiol were measured using RIQAS Chemiluminescent Immunoassay on Beckman Coulter Unicel Dxl 800 (16). Oestradiol measurements below the reportable range (175pmol/L) were considered the referent group. In the continuous analysis measurements below the reportable range were set to the cut-off value.

### Statistical analysis

Multivariable-adjusted logistic regression models estimated odds ratios for the presence of asthma associated with increasingly frequent shift work categories using day workers as the referent group.

We used three models, as in our prior study (15), including the following covariates: **Model 1** adjusted for age only; **Model 2** adjusted for age, ethnicity, chronotype, Townsend Deprivation Index (TDI), days exercised, alcohol status (current, previous or never), alcohol weekly intake, length of working week and whether the current job is considered to have an occupational asthma risk or requires a medical examination; **Model 3** adjusted for model 2 covariates plus potential mediators of the shift work-asthma relationship: BMI, smoking history, pack years smoked and sleep duration (17–22). Participants with missing covariate data were excluded from analyses.

Interaction analyses were performed by a likelihood-ratio test between models with/without interaction terms.

In all statistical tests p<0.05 was considered statistically significant. All analyses were performed in RStudio (v2023.12.1+402).

## Results

### Population characteristics

Demographic details by sex and shift work status are shown in **Table 1**. Male and female workers were similarly aged; females tended to smoke less than males. Female workers tended to drink less alcohol than males; both sexes showed similar trends across the work schedule groups. The ethnicity of participants was similar between the sexes. Female workers, especially those working nights, tended to work fewer weekly hours than males and come from more deprived areas.

**Table 1:**
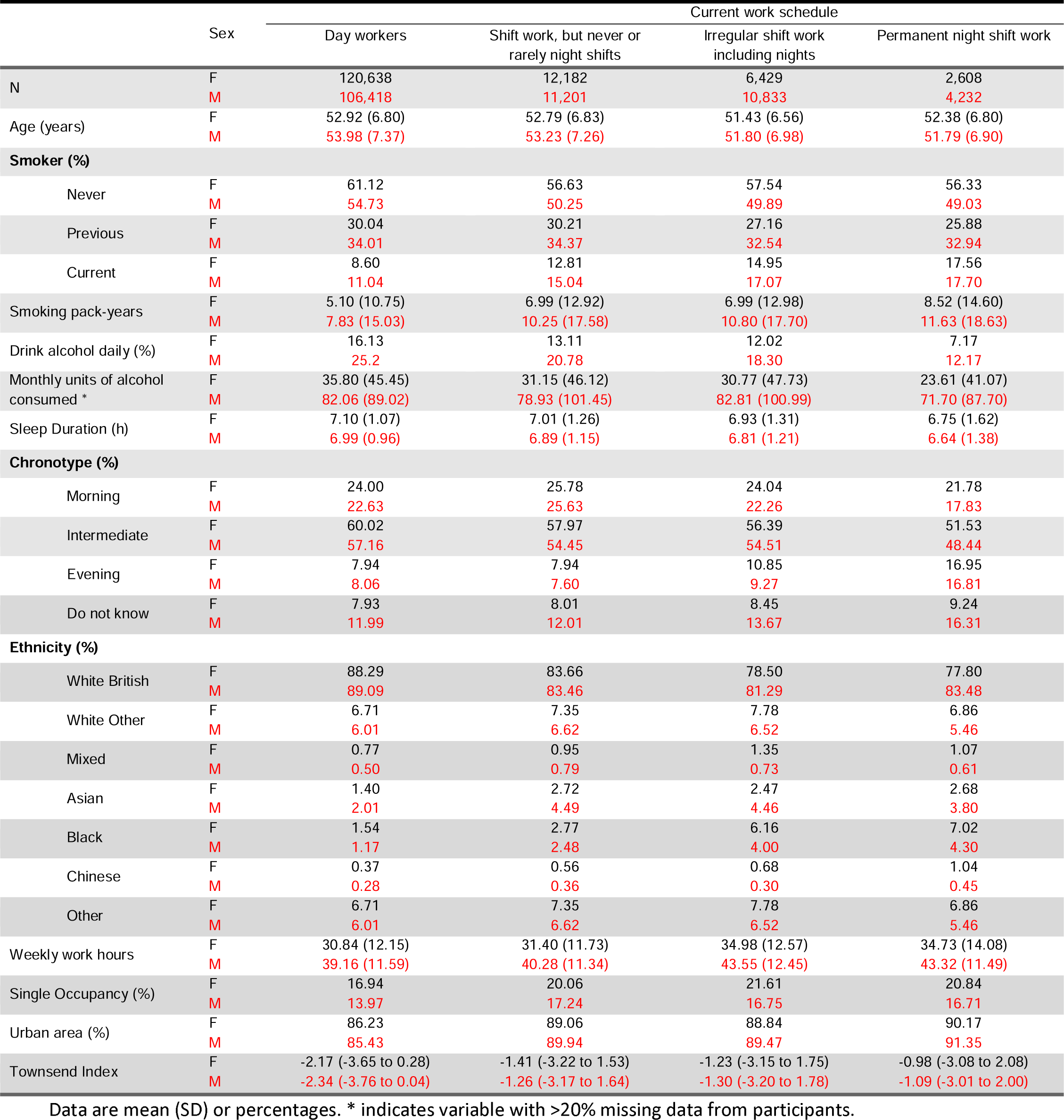
Social-demographic characteristics by current shift work exposure (n=274,541)

Health characteristics are presented in **Table 2**. Female and male workers showed similar trends in BMI with increasing BMI associated with greater frequency of shift work, especially in females.

**Table 2:** Health characteristics by current shift work exposure (n=274,541)

|  | Sex | Current work schedule |  |  |  |
| --- | --- | --- | --- | --- | --- |
|  |  | Day workers | Shift work, but never or rarely night shifts | Irregular shift work including nights | Permanent night shift work |
| N | F | 120,638 | 12,182 | 6,429 | 2,608 |
|  | M | 106,418 | 11,201 | 10,833 | 4,232 |
| BMI (kg/m^2) | F | 26.64 (5.05) | 27.42 (5.40) | 27.69 (5.59) | 28.47 (5.78) |
|  | M | 27.61 (4.06) | 28.20 (4.42) | 28.51 (4.39) | 28.54 (4.20) |
| Birth Weight (kg) | F | 3.25 (0.61) | 3.23 (0.66) | 3.24 (0.64) | 3.22 (0.69) |
|  | M | 3.45 (0.65) | 3.42 (0.69) | 3.44 (0.69) | 3.38 (0.72) |
| High Cholesterol (%) | F | 5.36 | 6.23 | 6.18 | 7.67 |
|  | M | 10.79 | 11.05 | 10.00 | 10.56 |
| Sleep Apnoea (%) | F | 0.11 | 0.10 | 0.20 | 0.04 |
|  | M | 0.48 | 0.55 | 0.54 | 0.40 |
| Chronic Obstructive Airways Disease/COPD | F | 0.15 | 0.21 | 0.19 | 0.19 |
|  | M | 0.12 | 0.21 | 0.19 | 0.09 |
| Emphysema/Chronic Bronchitis | F | 0.64 | 0.97 | 1.06 | 0.88 |
|  | M | 0.76 | 1.20 | 1.16 | 1.28 |
| Bronchiectasis (%) | F | 0.15 | 0.16 | 0.06 | 0.15 |
|  | M | 0.12 | 0.09 | 0.01 | 0.12 |
| Interstitial Lung Disease (%) | F | 0.01 | 0.01 | 0.00 | 0.00 |
|  | M | 0.03 | 0.01 | 0.05 | 0.02 |
| Other Respiratory Problems (%) | F | 0.11 | 0.13 | 0.14 | 0.12 |
|  | M | 0.12 | 0.21 | 0.16 | 0.05 |
| Gastro-Oesophageal Reflux (%) | F | 2.83 | 3.44 | 3.42 | 4.26 |
|  | M | 3.63 | 3.96 | 4.17 | 4.23 |
| Type 1 Diabetes (%) | F | 0.06 | 0.07 | 0.03 | 0.08 |
|  | M | 0.08 | 0.05 | 0.05 | 0.05 |
| Type 2 Diabetes (%) | F | 0.28 | 0.42 | 0.39 | 0.42 |
|  | M | 0.57 | 0.77 | 0.71 | 0.66 |
| Hypertension (%) | F | 17.21 | 19.14 | 18.34 | 20.28 |
|  | M | 23.80 | 25.60 | 24.24 | 25.17 |
| Cardiovascular Disease (%) | F | 2.20 | 2.94 | 2.80 | 2.91 |
|  | M | 5.65 | 5.93 | 5.21 | 4.87 |
| Depression (%) | F | 5.97 | 7.31 | 7.11 | 7.36 |
|  | M | 3.13 | 3.52 | 3.07 | 3.26 |
| Anxiety/panic attacks (%) | F | 1.39 | 1.53 | 1.28 | 1.30 |
|  | M | 0.89 | 0.95 | 0.66 | 0.76 |
| Hayfever/Allergic Rhinitis/Allergy to house dust mite (%) | F | 6.31 | 5.82 | 5.62 | 5.21 |
|  | M | 6.83 | 5.76 | 5.86 | 4.96 |
| Eczema/Dermatitis (%) | F | 2.75 | 2.35 | 2.77 | 2.34 |
|  | M | 2.69 | 2.42 | 2.10 | 2.08 |
| Allergy or Anaphylactic reaction to food/drug (%) | F | 1.57 | 1.48 | 1.62 | 1.73 |
|  | M | 0.88 | 0.82 | 0.81 | 0.73 |
| Testosterone (nmol/L) | F | 1.15 (0.68) | 1.14 (0.68) | 1.16 (0.67) | 1.19 (1.05) |
|  | M | 12.06 (3.62) | 12.11 (3.79) | 12.10 (3.65) | 12.31 (3.82) |
| Oestradiol (pmol/L) | F | 551.72 (487.86) | 539.76 (425.59) | 541.74 (453.25) | 551.5 (448.08) |
|  | M | 226.06 (88.75) | 225.06 (70.61) | 223.48 (79.49) | 229.42 (77.60) |
| SHBG (nmol/L) | F | 63.76 (31.78) | 62.00 (32.50) | 61.61 (31.80) | 62.11 (32.24) |
|  | M | 37.44 (15.59) | 36.91 (16.11) | 35.99 (15.31) | 36.51 (15.83) |
| Free Androgen Index | F | 2.34 (2.30) | 2.43 (2.37) | 2.48 (2.89) | 2.51 (2.93) |
|  | M | 35.57 (14.24) | 36.56 (19.71) | 37.40 (19.28) | 37.70 (17.35) |
| Free Testosterone (nmol/L) | F | 0.02 (0.01) | 0.02 (0.01) | 0.02 (0.01) | 0.02 (0.02) |
|  | M | 0.23 (0.07) | 0.23 (0.07) | 0.23 (0.07) | 0.24 (0.07) |

Data are mean (SD) or percentages.

### Female shift workers have higher odds of moderate-severe asthma

We first investigated sex differences in the association between increasing frequency of shift work and prevalent asthma. After adjusting for age (model 1), female shift workers had higher odds of moderate-severe asthma than female day workers (**Supplementary table 1:** shift work, but never or rarely night shifts, OR 1.16 (95% CI 1.02 – 1.32); irregular shift work including nights, OR 1.18 (95% CI 0.99 – 1.40); permanent night shift work, OR 1.54 (95% CI 1.22 – 1.95)). In contrast, male shift workers did not have a higher likelihood of moderate-severe asthma when compared to male day workers (shift work, but never or rarely night shifts, OR 1.05 (95% CI 0.89 – 1.23); irregular shift work including nights, OR 0.94 (95% CI 0.79 – 1.13); permanent night shift work, OR 0.91 (95% CI 0.69 – 1.20)).

Some of these relationships were maintained, but were attenuated after covariate adjustment (**Figure 1A (left), Supplementary table 1;** model 2, e.g. female permanent night shift workers, OR 1.50 95% CI (1.18-1.91)) and after further adjustment for potential mediators (sleep duration, smoking and BMI; **Figure 1A (right);** model 3, e.g. female permanent night shift workers, OR 1.31 95% CI (1.03-1.67)). An interaction between sex and the frequency of shift work was found across all models, indicating that increasing frequency of shift work was more strongly related to a higher likelihood of prevalent moderate-severe asthma in females than in males (p = 0.01).

**Figure 1:**
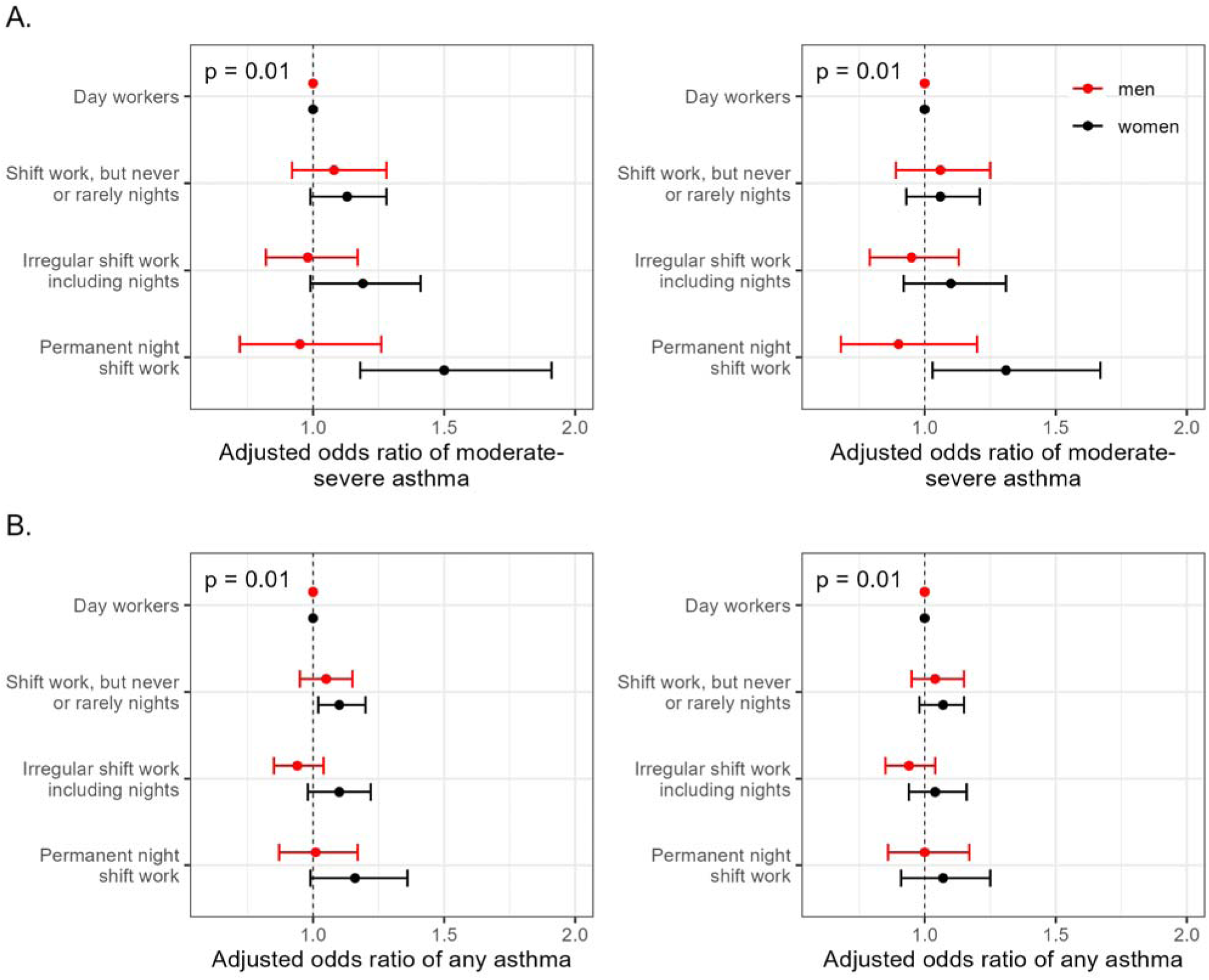
Adjusted odds (95% CI) of asthma by current shift work exposure, stratified by sex. Odds ratios of moderate-severe asthma (A; n=245,356; 117,350 male and 128,006 female) and any asthma (B; n=254,396; 121,387 male and 133,009 female). (Left) Adjusted for covariates in Model 2: age, ethnicity, Townsend deprivation index, alcohol status, daily alcohol intake, days exercised (walked, moderate and vigorous), length of working week, job asthma risk, job medical required and chronotype. (Right) Adjusted for covariates in Model 3: Model 2 covariates plus sleep duration, smoking status, smoking pack years and BMI.

Female shift workers also had higher odds of any asthma than female day workers (after adjusting for model 2 covariates: shift work, but never or rarely night shifts, OR 1.11 (95% CI 1.02 – 1.20); irregular shift work including nights, OR 1.10 (95% CI 0.99 – 1.22); permanent night shift work, OR 1.17 (95% CI 0.99 – 1.37)) whereas male shift workers showed no such corresponding relationships (shift work, but never or rarely night shifts, OR 1.05 (95% CI 0.95 – 1.15); irregular shift work including nights, OR 0.94 (95% CI 0.85 – 1.04); permanent night shift work, OR 1.00 (95% CI 0.86 – 1.17); **Supplementary table 2**). We found evidence of an interaction between sex and shift work status indicating that increasing frequency of shift work was more strongly related to the presence of ‘any asthma’ in females than in males (p = 0.01, **Figure 1B (left)**). The significant sex shift work interaction persisted in models adjusted for age (model 1; **Supplementary table 2**) and after adjusting for potential mediators (model 3; **Figure 1B (right)**)

Both female shift workers and male shift workers had a higher risks of experiencing wheeze or whistling in the chest (within the last year) than day workers; **Figure 2A (left)** (e.g. after adjusting for model 2 covariates; permanent female night shift workers OR 1.49 (95% CI 1.36 – 1.64) and male permanent night shift workers OR 1.31 (95% CI 1.22 – 1.41)). We found evidence of a sex-shift work interaction (p=0.01; model 2). The increased odds in male and female shift workers over day workers persisted in models 1 and 3, however a significant sex shift work interaction was only found in models 1 and 2 not in model 3 after controlling for potential mediators (**Figure 2A (right)**).

**Figure 2:**
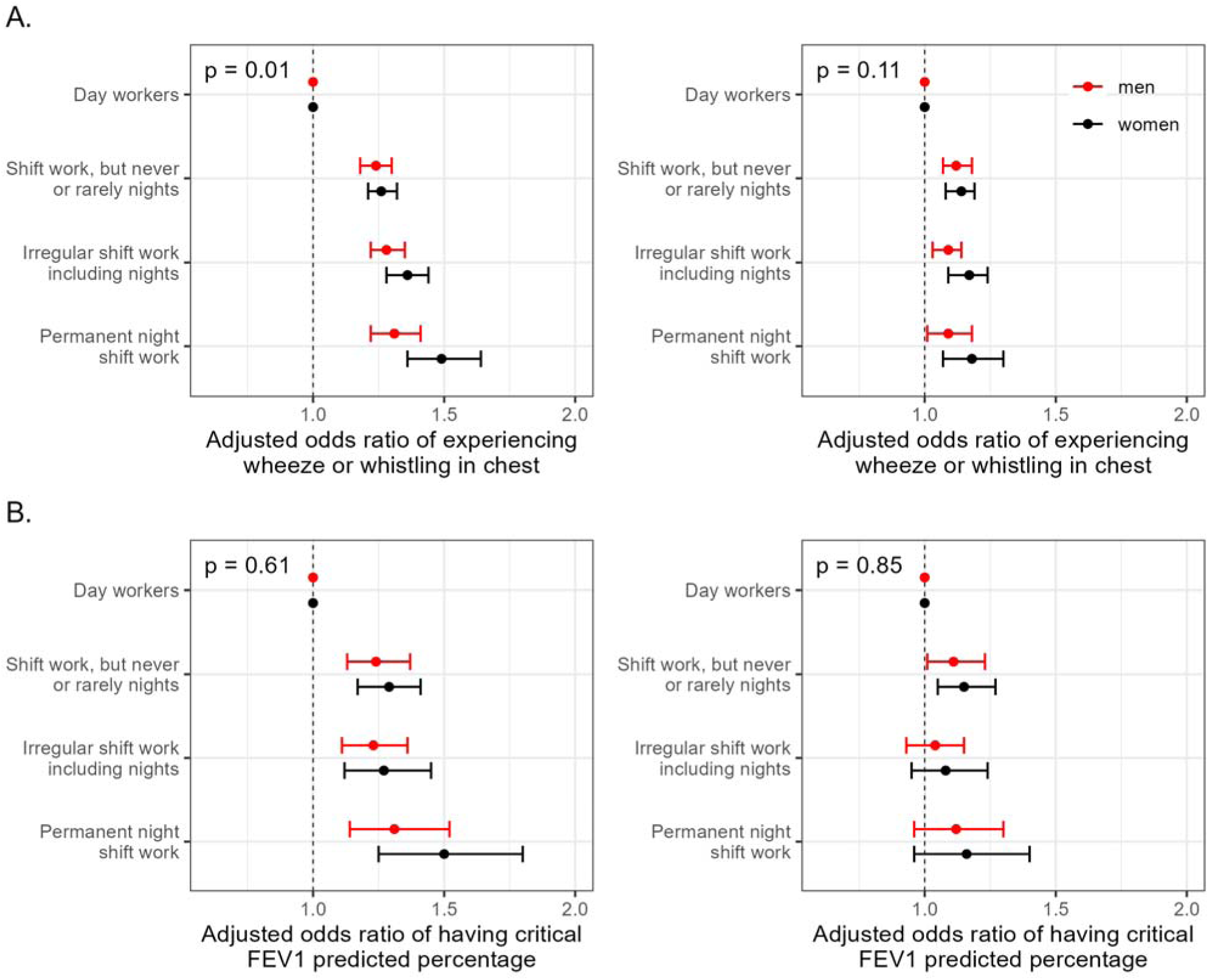
Adjusted odds (95% CI) of asthma symptoms by current shift work exposure, stratified by sex. Odds ratios of experiencing wheeze or whistling in chest within the last year (A; n=268,255; 127,374 male and 140,881 female) and of having a critical (<80%) FEV1 predicted percentage (B; n=87,358; 39,061 male and 48,297 female). (Left) Adjusted for covariates in Model 2: age, ethnicity, Townsend deprivation index, alcohol status, daily alcohol intake, days exercised (walked, moderate and vigorous), length of working week, job asthma risk, job medical required and chronotype. (Right) Adjusted for covariates in Model 3: Model 2 covariates plus sleep duration, smoking status, smoking pack years and BMI.

Further, female shift workers also had a higher risk of having obstructed lung function when compared to female day workers as did male shift workers when compared to male day workers; **Figure 2B (left)** (e.g. after adjusting for model 2 covariates permanent female night shift workers OR 1.50 (95% CI 1.25 – 1.80) and male permanent night shift workers OR 1.31 (95% CI 1.14 – 1.52)). The higher odds in male and female shift workers over day workers persisted in models 1, however was lost in all shift work schedules, except for “shift work, but never or rarely nights” where it was attenuated, after the addition of potential mediators in model 3 (**Figure 2B (right)**). No sex-shift work interaction was found in any of the models (**Supplementary table 4**).

### Role of chronotype and sex hormones in the sex-specific relationships linking shift work with prevalent asthma

We previously showed that moderate-severe asthma was linked to extreme chronotype (15). These associations remained in sex-stratified analyses (**Figure 3A**) with no evidence of an interaction between sex and chronotype (**Supplementary table 5**). Inclusion of chronotype in sex-stratified models of shift work-asthma relationships had negligible effects compared to base models, including no change in the interaction between sex and shift work. (**Figure 3B, Supplementary table 6**).

**Figure 3:**
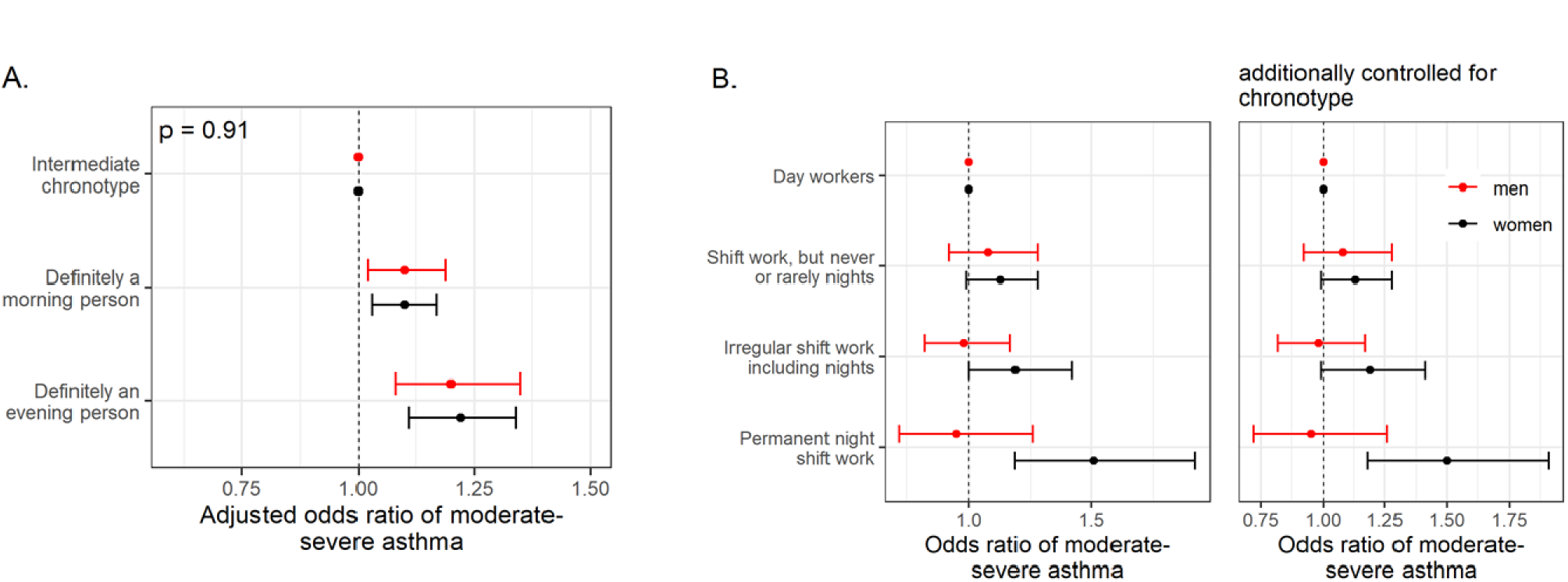
A: Adjusted odds (95% CI) of moderate-severe asthma by chronotype, stratified by sex. (n=377,515; 165,629 male and 211,886 female, model 2). B: Adjusted odds (95% CI) of moderate-severe asthma by current shift work exposure stratified by sex. Odds ratios of moderate-severe asthma from model without chronotype (left) and model including chronotype (right) (n=245,356; 117,350 male and 128,006 female, model 2). Model 2 covariates: age, ethnicity, Townsend deprivation index, alcohol status, daily alcohol intake, days exercised (walked, moderate and vigorous), length of working week, job asthma risk, job medical required.

Higher testosterone and SHBG were associated with lower prevalences of moderate-severe asthma in both females and males (**Supplementary figures 1A and 1B**; **Supplementary tables 7-8**). Corresponding relationships with oestradiol were null (**Supplementary figure 1C, Supplementary table 9**). Adding testosterone, SHBG or oestradiol into sex-stratified models of shift work-asthma relationships had negligible effects when compared to base models (**Supplementary figure 3, Supplementary tables 10-12**).

These data suggest that chronotype, testosterone, SHBG and oestradiol do not have major roles in moderating/mediating sex differences observed in relationships between shift work and prevalent asthma.

### Menopause and asthma

When compared to premenopausal females, postmenopausal females had a similar likelihood of having moderate-severe asthma (**Figure 4A, Supplementary table 13**). However, females who were unsure of their menopausal status and reported having had a hysterectomy, with/without an oophorectomy, had 1.4-1.5 fold higher adjusted odds of moderate-severe asthma when compared to premenopausal females **(Supplementary table 13**). These results attenuated to the null after excluding females using exogenous sex hormones, such as oral contraceptive pill (OCP) or hormone replacement therapy (HRT) (**Figure 4B, Supplementary table 14**).

**Figure 4:**
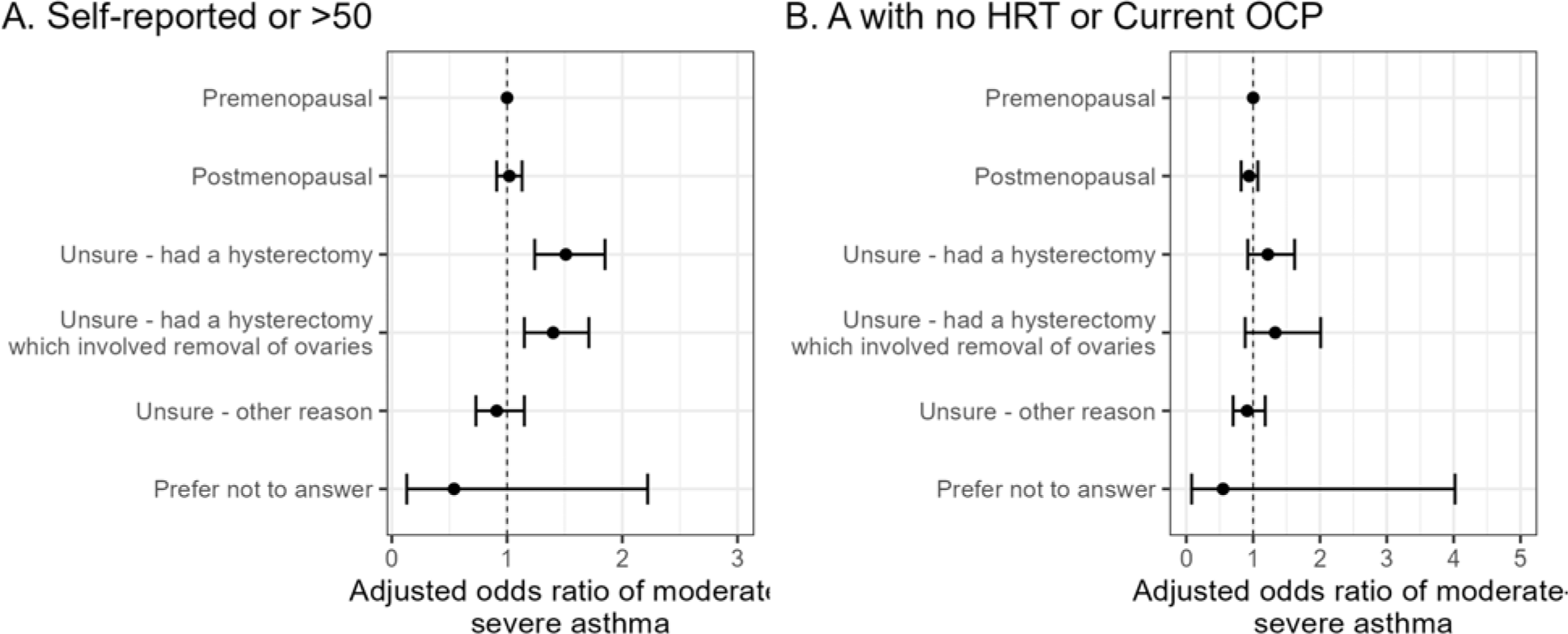
Adjusted odds (95% CI) of moderate-severe asthma by menopause status. A: Odds ratios of moderate-severe asthma by self-reported menopause status (postmenopausal status additionally include females > 50) (n=221,331). B: Odds ratios of moderate-severe asthma after excluding females using exogenous sex hormones, such as oral contraceptive pill (OCP) or hormone replacement therapy (HRT) (n=139,822). Adjusted by Model 2 covariates: age, ethnicity, Townsend deprivation index, alcohol status, daily alcohol intake, days exercised (walked, moderate and vigorous), length of working week, job asthma risk, job medical required.

As a sensitivity analysis we reran the shift work moderate-severe asthma analysis in females who had not had a hysterectomy (**Supplementary table 15**); obtaining similar results to those observed in females previously (**Figure 1A**).

### Role of menopause in the relationship between shift work and prevalent asthma

When compared to premenopausal day workers, premenopausal permanent night shift-workers and premenopausal females who worked “shift work, but never or rarely nights” had higher likelihoods of moderate-severe asthma (**Figure 5A**), but these relationships attenuated to the null after adjusting for potential mediators (**Supplementary table 16**). No relationships were found in postmenopausal/hysterectomy groups. We found no statistically significant interactions between menopause/hysterectomy status and the shift work-asthma relationship in any models (**Supplementary table 16**).

**Figure 5:**
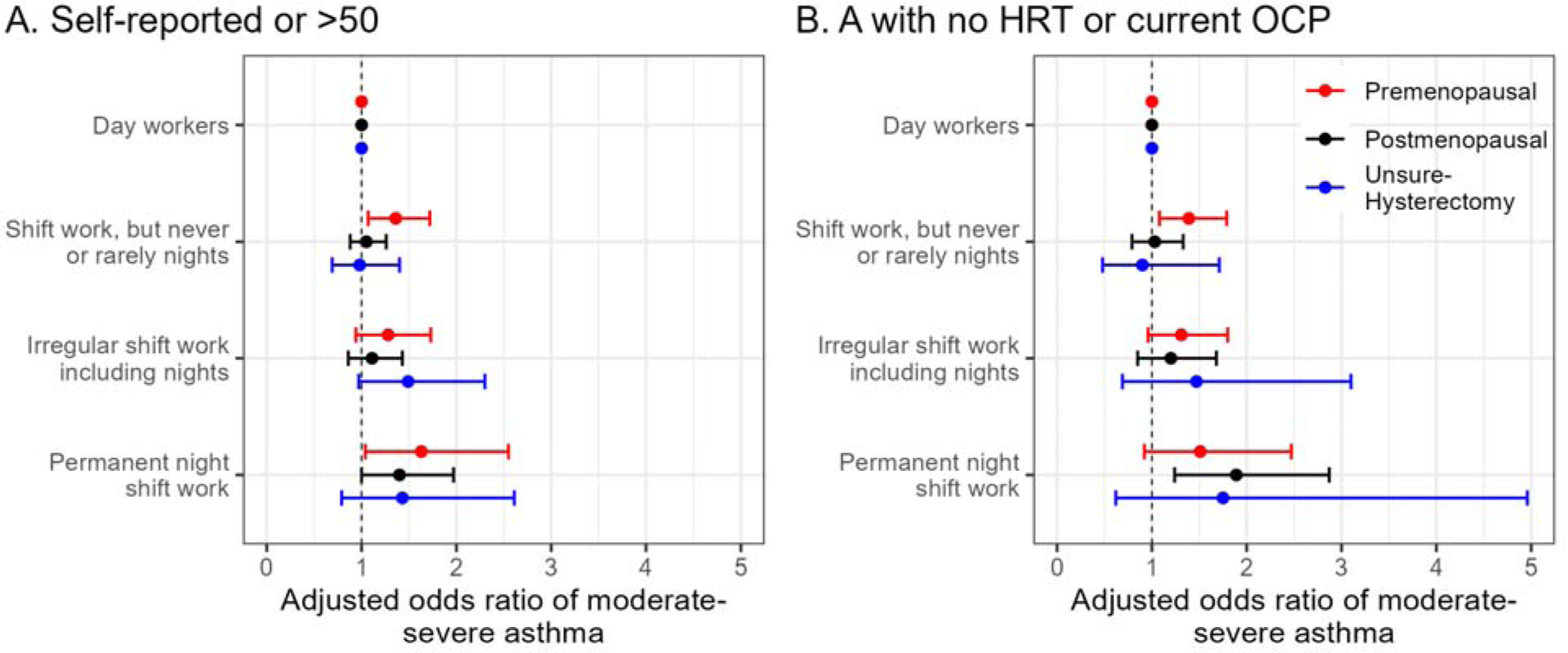
Adjusted odds (95% CI) of moderate-severe asthma by current shift work exposure, stratified by menopause status. A: Odds ratios of moderate-severe asthma by self-reported menopause status (postmenopausal status additionally include females > 50) (n=124,374). B: Odds ratios of moderate-severe asthma after excluding females using exogenous sex hormones, such as oral contraceptive pill (OCP) or hormone replacement therapy (HRT) (n=86,485). Adjusted by Model 2 covariates: age, ethnicity, Townsend deprivation index, alcohol status, daily alcohol intake, days exercised (walked, moderate and vigorous), length of working week, job asthma risk, job medical required.

After excluding participants receiving HRT/OCP, premenopausal shift workers who never or rarely worked nights had a higher likelihood of moderate-severe asthma, but these relationships attenuated to the null after adjusting for potential mediators (**Figure 5B, Supplementary table 17**). After excluding participants receiving HRT/OCP, postmenopausal permanent night shift-workers had 1.9-fold higher odds of moderate-severe asthma than day workers this was robust to covariate adjustment. However, we found no evidence of an interaction between menopause/hysterectomy status overall in the shift work-asthma relationships, however numbers were small in these subgroups (**Supplementary table 17**).

To determine whether the sex effect on the shift work-related asthma relationship (**Figure 1**) was driven by menopausal status we tested for a sex-shift work interaction. An interaction between sex and shift work frequency was found between males and premenopausal females but not postmenopausal females (**Supplementary tables 18**). However, after excluding females receiving HRT/OCP, an interaction between sex and shift work frequency was found in both premenopausal and postmenopausal females, this relationship remained significant after covariate adjustment in postmenopausal females (**Supplementary table 19**).

## Discussion

Our study has several novel findings. First, we found a significant sex interaction in the relationship between shift work frequency and prevalent moderate-severe asthma. Second, we showed that females, and not males, were at higher risk of moderate-severe asthma if they were night shift workers when compared to day workers. Third, we identified a significant sex interaction in relationships between shift work frequency and “any asthma” and “experiencing wheeze or whistling in the chest in the last year”. Fourth, we showed data suggesting that HRT may offer some protection from moderate-severe asthma in post-menopausal female permanent night shift-workers. Our findings have important implications for female shift workers and should be considered in asthma clinics and in Public Health Guidance.

To date there have been no previous studies investigating sex differences in the relationship between shift work and asthma.

An important potential mechanism that could explain the sex shift work interaction is differences in sex hormone levels. We demonstrated that higher levels of testosterone and SHBG are protective against asthma in both males and females, as shown previously (23). We showed no relationship between oestradiol and prevalent moderate-severe asthma. Adding sex hormones as covariates in our models had no impact on the sex-shift work interaction for the relationship with asthma suggesting a limited role of sex hormones in explaining observed sex differences.

However, there are some important caveats to consider in relation to our findings: a) there was no time-of-sampling data; or b) time of menstrual cycle data; and c) 80% of oestradiol levels were below the detectible level, possibly due to the age range of females studied. Such missing data reduced our statistical power.

We found that female participants who had had a hysterectomy ± oophorectomy had a higher likelihood of moderate-severe asthma when compared to premenopausal females. This has been reported previously (24), and it may be that these procedures are acting as a marker for shorter lifetime exposure to oestrogen (24).

We showed that postmenopausal and premenopausal permanent night shift-workers had higher odds of moderate-severe asthma when compared to corresponding day workers, but only premenopausal females had a significant interaction between sex and shift work frequency. However, when postmenopausal females taking HRT or OCP were excluded, female postmenopausal permanent night shift workers had 1.9-fold higher odds of moderate-severe asthma when compared to female day workers; and a significant sex interaction was observed across all models.

Together this suggests that HRT or OCP use in post-menopausal females could be protective for moderate-severe asthma in the context of shift work.

Previous studies investigating the effect of menopause on asthma have come to different conclusions; some suggesting that the menopause is protective for asthma (25, 26) and others showing the opposite (27). The role of HRT in asthma is also currently unclear; several observational studies have suggested that HRT might have adverse effects on asthma risk (24–26, 28). However, the largest prospective study performed to date showed that previous/current HRT use was associated with 17-21% lower risks of developing asthma (29). Other recent data suggests that the type of HRT might be important in determining whether HRT is a risk for, or protective against asthma (30).

Our results would suggest that, in postmenopausal shift workers, HRT might be protective for nightshift work-related asthma, however further research is needed to test this hypothesis in prospective studies and randomised controlled trials (RCT). These studies could test whether HRT protects against the development of asthma or reduces the severity of asthma in both dayworkers and in shift workers. It is currently unclear if shift work-related asthma risk would respond differently to HRT than asthma risk in day workers.

Previous studies have shown that the highest risk of HRT is in people using conjugated oestrogens (25, 31), who have a low BMI (28, 32) and are non-smokers (33). Our results also suggest that BMI and smoking moderate the relationship between sex and asthma.

We have shown previously that having an extreme chronotype (i.e. a definite preference for mornings or evenings) is associated with prevalent asthma (15). We know that males and females have differing distributions of chronotype with women tending to have earlier chronotypes, providing a potential mechanism for the sex differences observed. However, we found higher likelihood of moderate-severe asthma in extreme chronotypes in both male and females, and no evidence of a sex-chronotype interaction. Including chronotype in models linking shift work to moderate-severe asthma made no difference to results in either males or females. We conclude that chronotype does not explain the sex differences observed in the shift work asthma relationships.

Attenuation in our models suggests some effect of potential mediator variables on the sex differences observed. These variables, smoking, sleep duration and BMI, differed between the sexes with females having a higher increases of BMI and proportion of current smokers in shift work groups as compared to day workers. However, these mediators did not account for all of the effect observed as sex-shift work interactions stayed significant in Model 3.

Our study has several strengths. First, this is the first study to evaluate sex differences in the relationship between shift work frequency and asthma; an important topic from a public health perspective. Second, we do this in a large cohort (n=502,540) of individuals with linked health and socio-economic variables. Third, we employed a robust definition of moderate-severe asthma (15), likely capturing those with active disease. Fourth, we employed a staged modelling strategy accounting for important confounders and included potential mediators. These models accounted for the effects of risk factors that a) showed sex differences; b) were linked to shift work; and c) could have a causal role in asthma. Finally, we explored potential mechanisms explaining how our results linked to differing chrontypes and sex hormone levels.

Our study has some limitations. First, residual confounding could affect our results. For example, undefined occupational exposures could have effects on prevalent asthma especially when considering the different types of work undertaken by males and females (**Supplementary table 20**). However, we took steps to mitigate for this effect by including covariates that indicated jobs with high asthma risk or where a medical assessment for asthma was undertaken prior to employment. Second, in some subgroups of participants sample sizes were small resulting in limited power. Third, causal inference in a cross sectional study is not possible. Fourth, we potentially underestimate the detrimental effects of shift work on asthma caused by people becoming unwell as a result of their shift work and leaving employment. Lastly, the generalizability of our results may be limited as UK Biobank participants are generally healthier than the background population and have limited age, ethnic and social diversity (15).

Despite these limitations, our findings could have significant public health and clinical implications. The ability to reduce the risk of females developing asthma through modification of work schedules or through the introduction of HRT at menopause could have significant health-economic benefits.

### Interpretation

Our study shows that the higher likelihood of moderate-severe asthma in shift workers compared to day workers is present in females but not in males. The mechanisms are unclear, but HRT may have a protective role in postmenopausal female shift workers. Future large RCTs are needed to test this hypothesis and provide clarity to an increasingly large proportion of our workforce.

## Data Availability

All data is available after request from the UK Biobank

https://www.ukbiobank.ac.uk/

## Abbreviations

BMI: Body Mass Index
CI: Confidence Interval
FEV1: Forced Expiratory Volume in 1 second
HRT: Hormone Replacement Therapy
OCP: Oral Contraceptive Pill
OR: Odds Ratio
RCT: Randomized Controlled Trial
SHBG: Sex Hormone Binding Globulin
TDI: Townsend Deprivation Index.

## Take-Home Points

### Study question

Is increasing frequency of night shift work more strongly related to prevalent asthma in females than in males?

### Results

Compared to female day workers, female permanent night shift workers had higher covariate-adjusted odds of moderate-severe asthma (OR: 1.50 (95% CI 1.18 – 1.91)) but there was no corresponding relationship in males (OR 0.95 (95% CI 0.72 – 1.26); sex interaction p-value = 0.01).

### Interpretation

Our finding that increasing night shift work frequency is more strongly related to asthma in females than in males could have Public Health implications.

