## Supplemental Figures and Tables for "Increased risk of asthma in female night shift workers"

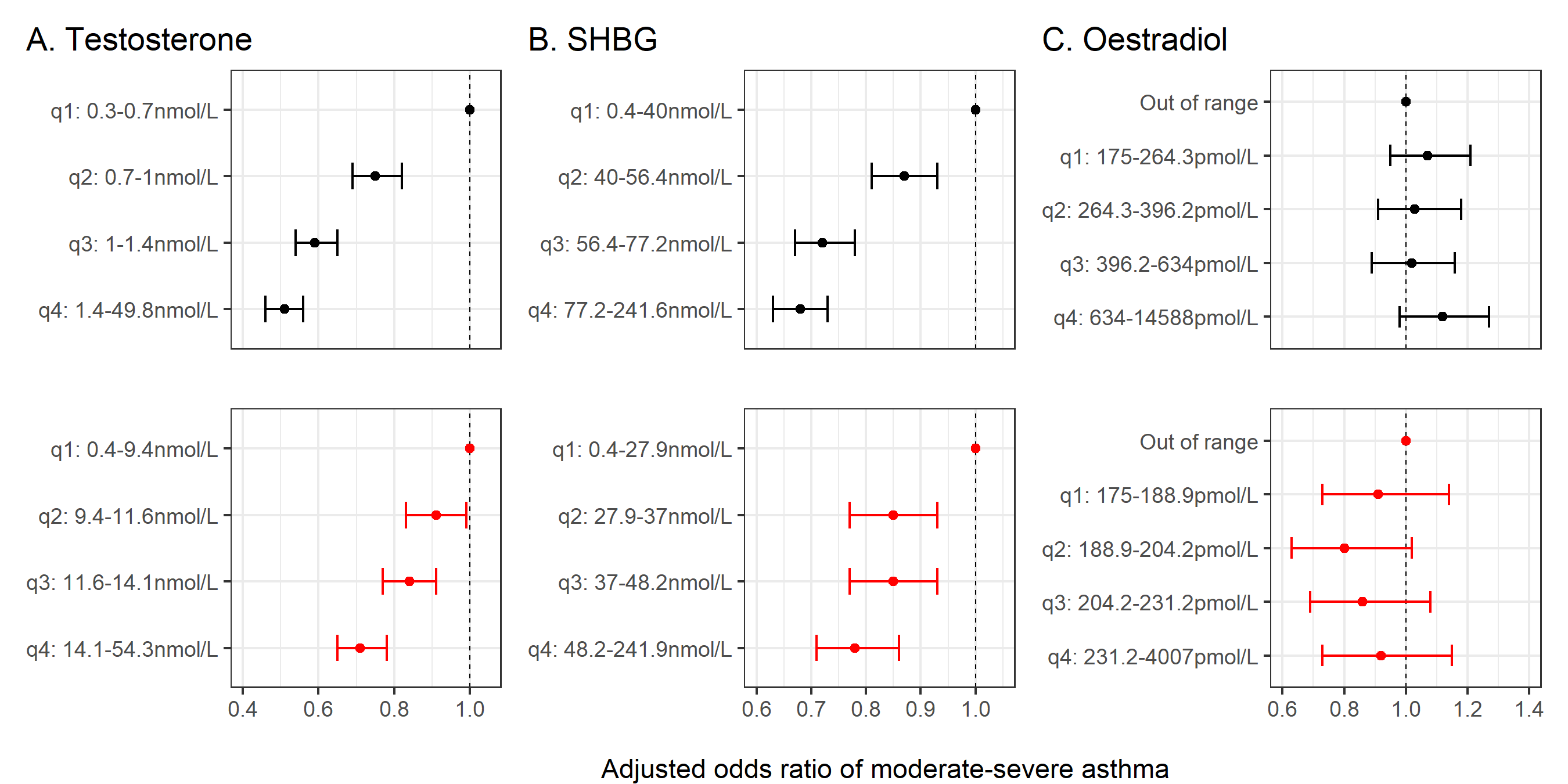

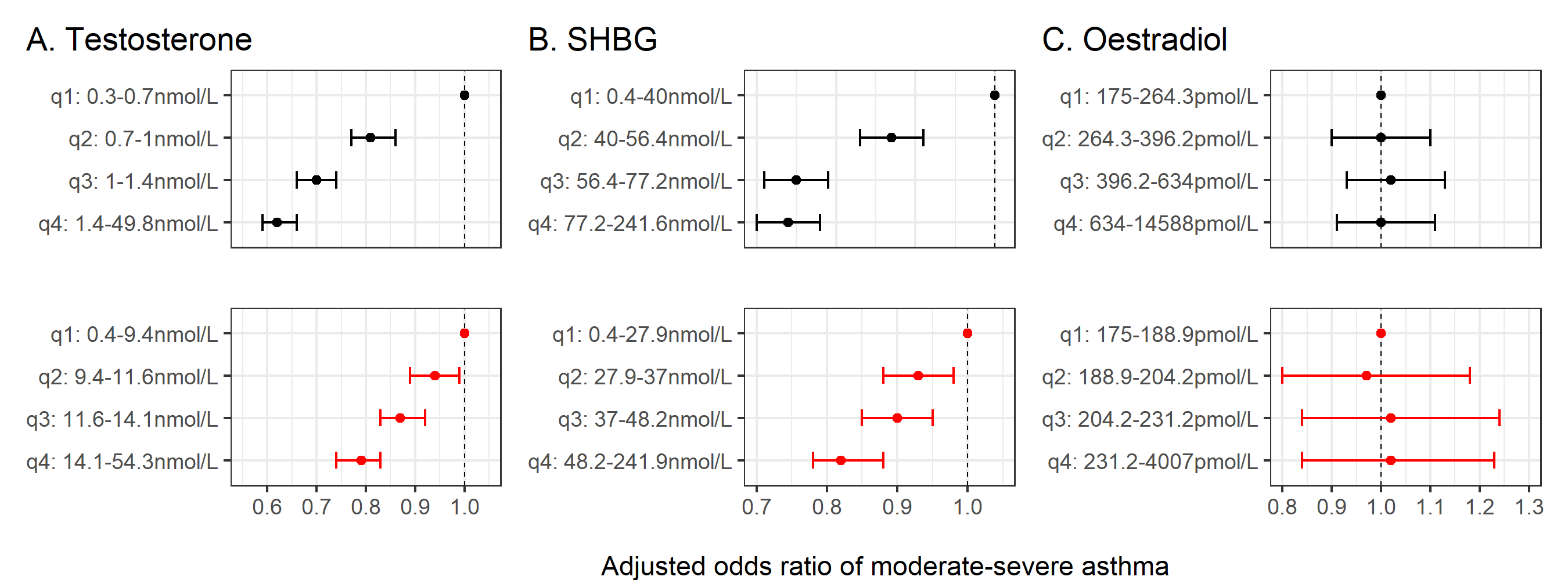

**Supplementary figure 1. Effect of sex hormones on moderate-severe asthma, model 2. Female (top), male (bottom). Adjusted for covariates in Model 2: age, ethnicity, Townsend deprivation index, alcohol status, daily alcohol intake, days exercised (walked, moderate and vigorous), length of working week, job asthma risk, job medical required and chronotype.**

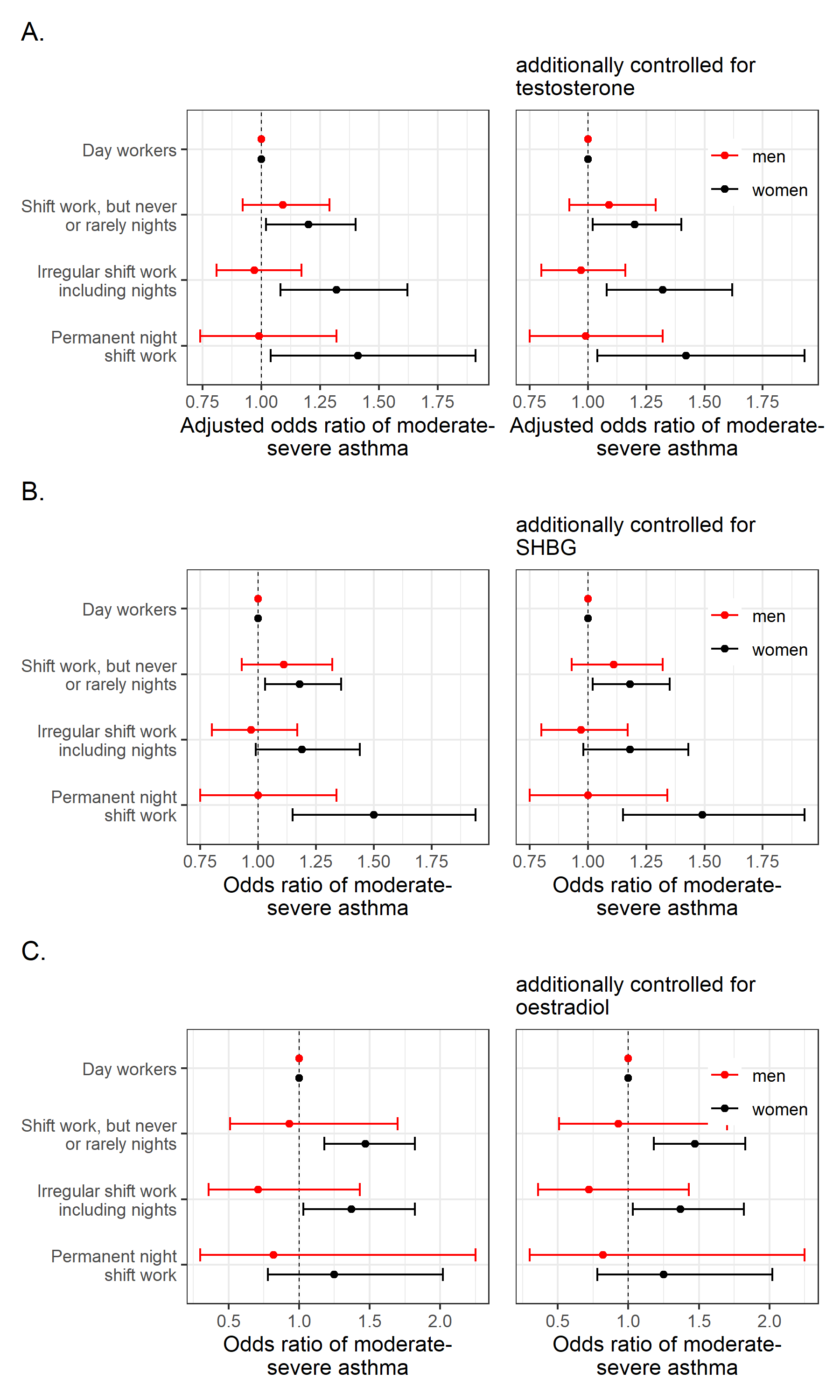

**Supplementary figure 2: Adjusted odds (95% CI) of moderate-severe asthma by current shift work exposure stratified by sex. Odds ratios of moderate-severe asthma from model 2 (left) and model 2 plus additionally controlled for an additional covariate (right). Covariates additionally controlled for; testosterone (A, n=216,364), SHBG (B, n=211,866) and oestradiol (C, n=49,540). Model 2 covariates: age, ethnicity, Townsend deprivation index, alcohol status, daily alcohol intake, days exercised (walked, moderate and vigorous), length of working week, job asthma risk, job medical required and chronotype.**

**Supplementary table 1: Adjusted odds (95% CI) of moderate-severe asthma by current shift work exposure with day workers as referent, stratified by sex. (n=245,356)**

| Female/Male |  | Current work schedule | | | Sex-shift work interaction |
| --- | --- | --- | --- | --- | --- |
|  | Day workers | Shift work, but never or rarely night shifts | Irregular shift work including nights | Permanent night shift work |  |
| Total cases (% of total sample size) | 2,254 (2.07%)  1,469 (1.56%) | 262 (2.39%)  161 (1.62%) | 139 (2.38%)  139 (1.43%) | 73 (3.13%)  52 (1.38%) |  |
| Total sample size | 108,849  93,937 | 10,983  9,941 | 5,841  9,709 | 2,333  3,763 |  |
| Model 1: Age-adjusted | Female referent  Male referent | **1.16 (1.02-1.32)**  1.05 (0.89-1.23) | 1.18 (0.99-1.40)  0.94 (0.79-1.13) | **1.54 (1.22-1.95)**  0.91 (0.69-1.20) | **0.01** |
| Model 2: Multivariable-adjusted | Female referent  Male referent | 1.13 (0.99-1.28)  1.08 (0.92-1.28) | 1.19 (0.99-1.41)  0.98 (0.82-1.17) | **1.50 (1.18-1.91)**  0.95 (0.72-1.26) | **0.01** |
| Model 3: Model 2 covariates + potential mediators | Female referent  Male referent | 1.06 (0.93-1.21)  1.06 (0.89-1.25) | 1.10 (0.92-1.31)  0.95 (0.79-1.13) | **1.31 (1.03-1.67)**  0.90 (0.68-1.20) | **0.01** |

Model 2 covariates: age, ethnicity, Townsend deprivation index, alcohol status, daily alcohol intake, days exercised (walked, moderate and vigorous), length of working week, chronotype, job asthma risk and job medical required. Model 3 data are adjusted for Model 2 covariates plus potential mediators sleep duration, smoking status, smoking pack years and BMI.

**Supplementary table 2: Adjusted odds (95% CI) of any asthma by current shift work exposure with day workers as referent, stratified by sex. (n=254,396)**

| Female/Male |  | Current work schedule | | | Sex-shift work interaction |
| --- | --- | --- | --- | --- | --- |
|  | Day workers | Shift work, but never or rarely night shifts | Irregular shift work including nights | Permanent night shift work |  |
| Total cases (% of total sample size) | 6,458 (5.71%)  4,715 (4.85%) | 730 (6.37%)  508 (4.94%) | 380 (6.25%)  450 (4.49%) | 163 (6.73%)  185 (4.75%) |  |
| Total sample size | 113,053  97,183 | 11,451  10,288 | 6,082  10,020 | 2,423  3,896 |  |
| Model 1: Age-adjusted | Female referent  Male referent | **1.12 (1.04-1.22)**  1.01 (0.92-1.11) | 1.09 (0.98-1.22)  **0.89 (0.81-0.99)** | **1.19 (1.01-1.40)**  0.95 (0.81-1.10) | **0.01** |
| Model 2: Multivariable-adjusted | Female referent  Male referent | **1.11 (1.02-1.20)**  1.05 (0.95-1.15) | 1.10 (0.99-1.22)  0.94 (0.85-1.04) | 1.17 (0.99-1.37)  1.00 (0.86-1.17) | **0.01** |
| Model 3: Model 2 covariates +  potential mediators | Female referent  Male referent | 1.06 (0.98-1.15)  1.04 (0.95-1.14) | 1.04 (0.93-1.16)  0.93 (0.84-1.03) | 1.07 (0.90-1.26)  0.99 (0.85-1.16) | **0.01** |

Model 2 covariates: age, ethnicity, Townsend deprivation index, alcohol status, daily alcohol intake, days exercised (walked, moderate and vigorous), length of working week, chronotype, job asthma risk and job medical required. Model 3 data are adjusted for Model 2 covariates plus potential mediators sleep duration, smoking status, smoking pack years and BMI.

**Supplementary table 3: Adjusted odds (95% CI) of experiencing wheeze or whistling in the chest within the last year by current shift work exposure with day workers as referent, stratified by sex. (n=268,255)**

| Female/Male |  | Current work schedule | | | Sex-shift work interaction |
| --- | --- | --- | --- | --- | --- |
|  | Day workers | Shift work, but never or rarely night shifts | Irregular shift work including nights | Permanent night shift work |  |
| Total cases (% of total sample size) | 20,666 (17.25%)  19,936 (19.47%) | 2,599 (21.50%)  2,503 (23.53%) | 1,449 (22.55%)  2,501 (24.23%) | 643 (25.10%)  993 (24.83%) |  |
| Total sample size | 119,807  102,414 | 12,087  10,639 | 6,425  10,322 | 2,562  3,999 |  |
| Model 1: Age-adjusted | Female referent  Male referent | **1.31 (1.26-1.38)**  **1.27 (1.21-1.33)** | **1.41 (1.33-1.50)**  **1.31 (1.25-1.38)** | **1.61 (1.47-1.77)**  **1.36 (1.26-1.46)** | **0.03** |
| Model 2: Multivariable-adjusted | Female referent  Male referent | **1.26 (1.21-1.32)**  **1.24 (1.18-1.30)** | **1.36 (1.28-1.44)**  **1.28 (1.22-1.35)** | **1.49 (1.36-1.64)**  **1.31 (1.22-1.41)** | **0.01** |
| Model 3: Model 2 covariates + potential mediators | Female referent  Male referent | **1.14 (1.08-1.19)**  **1.12 (1.07-1.18)** | **1.17 (1.09-1.24)**  **1.09 (1.03-1.14)** | **1.18 (1.07-1.30)**  **1.09 (1.01-1.18)** | 0.11 |

Model 2 covariates: age, ethnicity, Townsend deprivation index, alcohol status, daily alcohol intake, days exercised (walked, moderate and vigorous), length of working week, chronotype, job asthma risk and job medical required. Model 3 data are adjusted for Model 2 covariates plus potential mediators sleep duration, smoking status, smoking pack years and BMI.

**Supplementary table 4: Adjusted odds (95% CI) of having a critical (<80%) FEV1 predicted percentage by current shift work exposure with day workers as referent, stratified by sex. (n=88,950)**

| Female/Male |  | Current work schedule | | | Sex-shift work interaction |
| --- | --- | --- | --- | --- | --- |
|  | Day workers | Shift work, but never or rarely night shifts | Irregular shift work including nights | Permanent night shift work |  |
| Total cases (% of total sample size) | 4,800 (11.67%)  4,368 (14.05%) | 615 (15.04%)  547 (16.93%) | 303 (13.89%)  529 (15.83%) | 151 (17.00%)  237 (16.98%) |  |
| Total sample size | 41,140  31,094 | 4,088  3,230 | 2,181  3,341 | 888  1,396 |  |
| Model 1: Age-adjusted | Female referent  Male referent | **1.34 (1.23-1.47)**  **1.29 (1.17-1.42)** | **1.31 (1.15-1.48)**  **1.26 (1.14-1.39)** | **1.60 (1.34-1.91)**  **1.36 (1.18-1.57)** | 0.52 |
| Model 2: Multivariable-adjusted | Female referent  Male referent | **1.29 (1.17-1.41)**  **1.24 (1.13-1.37)** | **1.27 (1.12-1.45)**  **1.23 (1.11-1.36)** | **1.50 (1.25-1.80)**  **1.31 (1.14-1.52)** | 0.61 |
| Model 3: Model 2 covariates + potential mediators | Female referent  Male referent | **1.15 (1.05-1.27)**  **1.11 (1.01-1.23)** | 1.08 (0.95-1.24)  1.04 (0.93-1.15) | 1.16 (0.96-1.40)  1.12 (0.96-1.30) | 0.85 |

Model 2 covariates: age, ethnicity, Townsend deprivation index, alcohol status, daily alcohol intake, days exercised (walked, moderate and vigorous), length of working week, chronotype, job asthma risk and job medical required. Model 3 data are adjusted for Model 2 covariates plus potential mediators sleep duration, smoking status, smoking pack years and BMI.

**Supplementary table 5: Adjusted odds (95% CI) of moderate-severe asthma by chronotype with intermediate chronotype as referent, stratified by sex. (n=377,515)**

| Female/Male | Chronotype | | | Sex-chronotype interaction |
| --- | --- | --- | --- | --- |
|  | Intermediate | Definitely a morning person | Definitely an evening person |  |
| Total cases (% of total sample size) | 3,431 (2.53%)  2,101 (1.98%) | 1,665 (2.86%)  974 (2.20%) | 565 (3.18%)  373 (2.46%) |  |
| Total sample size | 135,840  106,135 | 58,290  44,312 | 17,756  15,182 |  |
| Model 1: Age-adjusted | Female referent  Male referent | **1.11 (1.05-1.18)**  **1.09 (1.01-1.17)** | **1.29 (1.18-1.41)**  **1.31 (1.17-1.47)** | 0.96 |
| Model 2: Multivariable-adjusted | Female referent  Male referent | **1.10 (1.03-1.17)**  **1.10 (1.02-1.19)** | **1.22 (1.11-1.34)**  **1.20 (1.08-1.35)** | 0.91 |
| Model 3: Model 2 covariates + potential mediators | Female referent  Male referent | **1.08 (1.02-1.15)**  1.08 (1.00-1.17) | **1.10 (1.00-1.21)**  **1.15 (1.03-1.29)** | 0.86 |

Model 2 covariates: age, ethnicity, Townsend deprivation index, alcohol status, daily alcohol intake, days exercised (walked, moderate and vigorous), length of working week, job asthma risk and job medical required. Model 3 data are adjusted for Model 2 covariates plus potential mediators sleep duration, smoking status, smoking pack years and BMI.

**Supplementary table 6: Adjusted odds (95% CI) of moderate-severe asthma by current shift work exposure with day workers as referent, stratified by sex. (n=245,356). Comparison of models with/without adjusting for chronotype.**

| Female/Male |  | Current work schedule | | | Sex-shift work interaction |
| --- | --- | --- | --- | --- | --- |
|  | Day workers | Shift work, but never or rarely night shifts | Irregular shift work including nights | Permanent night shift work |  |
| Total cases (% of total sample size) | 2,254 (2.07%)  1,469 (1.56%) | 262 (2.39%)  161 (1.62%) | 139 (2.38%)  139 (1.43%) | 73 (3.13%)  52 (1.38%) |  |
| Total sample size | 108,849  93,937 | 10,983  9,941 | 5,841  9,709 | 2,333  3,763 |  |
| Model 1: Age-adjusted | Female referent  Male referent | **1.16 (1.02-1.32)**  1.05 (0.89-1.23) | 1.18 (0.99-1.40)  0.94 (0.79-1.13) | **1.54 (1.22-1.95)**  0.91 (0.69-1.20) | **0.01** |
| Model 1 + chronotype | Female referent  Male referent | **1.16 (1.02-1.32)**  1.05 (0.89-1.24) | 1.18 (0.99-1.40)  0.94 (0.79-1.13) | **1.53 (1.21-1.94)**  0.91 (0.69-1.20) | **0.01** |
| Model 2 - chronotype | Female referent  Male referent | 1.13 (0.99-1.28)  1.08 (0.92-1.28) | 1.19 (1.00-1.42)  0.98 (0.82-1.17) | **1.51 (1.19-1.92)**  0.95 (0.72-1.26) | **0.01** |
| Model 2: Multivariable-adjusted | Female referent  Male referent | 1.13 (0.99-1.28)  1.08 (0.92-1.28) | 1.19 (0.99-1.41)  0.98 (0.82-1.17) | **1.50 (1.18-1.91)**  0.95 (0.72-1.26) | **0.01** |
| Model 3 - chronotype | Female referent  Male referent | 1.06 (0.93-1.21)  1.06 (0.89-1.25) | 1.10 (0.92-1.31)  0.95 (0.79-1.13) | **1.32 (1.03-1.68)**  0.90 (0.68-1.20) | **0.01** |
| Model 3: Model 2 covariates +  potential mediators | Female referent  Male referent | 1.06 (0.93-1.21)  1.06 (0.89-1.25) | 1.10 (0.92-1.31)  0.95 (0.79-1.13) | **1.31 (1.03-1.67)**  0.90 (0.68-1.20) | **0.01** |

Model 2 covariates: age, ethnicity, Townsend deprivation index, alcohol status, daily alcohol intake, days exercised (walked, moderate and vigorous), length of working week, chronotype, job asthma risk and job medical required. Model 3 data are adjusted for Model 2 covariates plus potential mediators sleep duration, smoking status, smoking pack years and BMI.

**Supplementary table 7: Adjusted odds (95% CI) of moderate-severe asthma by testosterone with first quartile of testosterone as referent, stratified by sex. (n=379,395)**

| Female  (n=189,029) | Testosterone quartile | | | | p-value  for trend |
| --- | --- | --- | --- | --- | --- |
|  | q1: 0.4-0.7nmol/L | q2: 0.7-1.0nmol/L | q3: 1.0-1.4nmol/L | q4: 1.4-49.8nmol/L |  |
| Total cases (% of total sample size) | 1,309 (2.80%) | 991 (2.10%) | 797 (1.68%) | 701 (1.47%) |  |
| Total sample size | 46,707 | 47,191 | 47,386 | 47,745 |  |
| Model 1: Age-adjusted OR (95% CI) | Female referent | **0.75 (0.69-0.81)** | **0.60 (0.55-0.66)** | **0.52 (0.48-0.58)** | **<0.01** |
| Model 2: Multivariable adjusted OR (95% CI) | Female referent | **0.75 (0.69-0.82)** | **0.59 (0.54-0.65)** | **0.51 (0.46-0.56)** | **<0.01** |
| Model 3: Model 2 covariates + potential mediators (95% CI) | Female referent | **0.73 (0.67-0.79)** | **0.56 (0.51-0.61)** | **0.45 (0.41-0.49)** | **<0.01** |
| Male  (n=190,366) | q1: 0.4-9.4nmol/L | q2: 9.4-11.6nmol/L | q3: 11.6-14.1nmol/L | q4: 14.1-54.3nmol/L |  |
| Total cases (% of total sample size) | 1,185 (2.51%) | 1,029 (2.16%) | 937 (1.96%) | 800 (1.67%) |  |
| Total sample size | 47,181 | 47,586 | 47,746 | 47,853 |  |
| Model 1: Age- adjusted OR (95% CI) | Male referent | **0.87 (0.80-0.95)** | **0.79 (0.73-0.87)** | **0.68 (0.62-0.75)** | **<0.01** |
| Model 2: Multivariable adjusted OR (95% CI) | Male referent | **0.91 (0.83-0.99)** | **0.84 (0.77-0.91)** | **0.71 (0.65-0.78)** | **<0.01** |
| Model 3: Model 2 covariates + potential mediators (95% CI) | Male referent | 0.96 (0.88-1.05) | 0.93 (0.85-1.02) | **0.80 (0.73-0.89)** | **<0.01** |

Model 2 covariates: age, ethnicity, Townsend deprivation index, alcohol status, daily alcohol intake, days exercised (walked, moderate and vigorous), length of working week, chronotype, job asthma risk and job medical required. Model 3 data are adjusted for Model 2 covariates plus potential mediators sleep duration, smoking status, smoking pack years and BMI.

**Supplementary table 8: Adjusted odds (95% CI) of moderate-severe asthma by SHBG with first quartile of SHBG as referent, stratified by sex. (n=378,342)**

| Female  (n=202,186) | SHBG quartile | | | | p-value  for trend |
| --- | --- | --- | --- | --- | --- |
|  | q1: 0.4-40.0nmol/L | q2: 40.0-56.4nmol/L | q3: 56.4-77.2nmol/L | q4: 77.2-241.6nmol/L |  |
| Total cases (% of total sample size) | 1,718 (3.45%) | 1,441 (2.85%) | 1,177 (2.31%) | 1,096 (2.15%) |  |
| Total sample size | 49,726 | 50,502 | 51,011 | 50,947 |  |
| Model 1: Age- adjusted OR (95% CI) | Female referent | **0.82 (0.76-0.88)** | **0.67 (0.62-0.72)** | **0.63 (0.58-0.68)** | **<0.01** |
| Model 2: Multivariable adjusted OR (95% CI) | Female referent | **0.87 (0.81-0.93)** | **0.72 (0.67-0.78)** | **0.68 (0.63-0.73)** | **<0.01** |
| Model 3: Model 2 covariates + potential mediators (95% CI) | Female referent | 1.01 (0.94-1.09) | 0.93 (0.86-1.01) | 0.96 (0.88-1.05) | **<0.01** |
| Male  (n=176,156) | q1: 0.4-27.9nmol/L | q2: 27.9-37.0nmol/L | q3: 37.0-48.2nmol/L | q4: 48.2-241.9nmol/L |  |
| Total cases (% of total sample size) | 964 (2.21%) | 876 (1.99%) | 917 (2.08%) | 912 (2.05%) |  |
| Total sample size | 43,555 | 44,001 | 44,141 | 44,459 |  |
| Model 1: Age- adjusted OR (95% CI) | Male referent | **0.83 (0.76-0.91)** | **0.83 (0.75-0.91)** | **0.78 (0.70-0.85)** | 0.68 |
| Model 2: Multivariable adjusted OR (95% CI) | Male referent | **0.85 (0.77-0.93)** | **0.85 (0.77-0.93)** | **0.78 (0.71-0.86)** | 0.81 |
| Model 3: Model 2 covariates + potential mediators (95% CI) | Male referent | 0.91 (0.82-1.00) | 0.91 (0.83-1.01) | **0.89 (0.80-0.99)** | 0.58 |

Model 2 covariates: age, ethnicity, Townsend deprivation index, alcohol status, daily alcohol intake, days exercised (walked, moderate and vigorous), length of working week, chronotype, job asthma risk and job medical required. Model 3 data are adjusted for Model 2 covariates plus potential mediators sleep duration, smoking status, smoking pack years and BMI.

**Supplementary table 9: Adjusted odds (95% CI) of moderate-severe asthma by oestradiol with participants recording measurements below the reportable range (<175 pmol/L) as referent, stratified by sex. (n=388,964)**

| Female  (n=209,700) | Oestradiol quartile | | | | | p-value  for trend |
| --- | --- | --- | --- | --- | --- | --- |
|  | Out of range  (too low) | q1: 175.0-264.3pmol/L | q2: 264.3-396.2pmol/L | q3: 396.2-634.0pmol/L | q4: 634.0-14,588pmol/L |  |
| Total cases (% of total sample size) | 4,410 (2.77%) | 316 (2.50%) | 286 (2.26%) | 272 (2.15%) | 298 (2.35%) |  |
| Total sample size | 159,076 | 12,630 | 12,667 | 12,634 | 12,693 |  |
| Model 1: Age- adjusted OR (95% CI) | Female referent | 1.09 (0.97-1.23) | 1.03 (0.91-1.18) | 1.01 (0.88-1.15) | 1.11 (0.97-1.26) | **<0.01** |
| Model 2: Multivariable adjusted OR (95% CI) | Female referent | 1.07 (0.95-1.21) | 1.03 (0.91-1.18) | 1.02 (0.89-1.16) | 1.12 (0.98-1.27) | **<0.01** |
| Model 3: Model 2 covariates + potential mediators (95% CI) | Female referent | 1.05 (0.93-1.19) | 1.04 (0.91-1.19) | 1.04 (0.91-1.20) | **1.19 (1.04-1.36)** | **<0.01** |
| Male  (n=179,264) | Out of range | q1: 175.0-188.9pmol/L | q2: 188.9-204.2pmol/L | q3: 204.2-231.2pmol/L | q4: 231.2-4,007pmol/L |  |
| Total cases (% of total sample size) | 3,426 (2.11%) | 82 (1.97%) | 71 (1.71%) | 78 (1.87%) | 82 (2.00%) |  |
| Total sample size | 162,683 | 4,161 | 4,140 | 4,173 | 4,107 |  |
| Model 1: Age-adjusted OR (95% CI) | Male referent | 0.93 (0.75-1.16) | 0.80 (0.63-1.02) | 0.88 (0.70-1.11) | 0.97 (0.77-1.21) | 0.30 |
| Model 2: Multivariable adjusted OR (95% CI) | Male referent | 0.91 (0.73-1.14) | 0.80 (0.63-1.02) | 0.86 (0.69-1.08) | 0.92 (0.73-1.15) | 0.34 |
| Model 3: Model 2 covariates + potential mediators (95% CI) | Male referent | 0.94 (0.75-1.18) | 0.81 (0.64-1.03) | 0.84 (0.66-1.06) | 0.91 (0.72-1.14) | 0.35 |

p-value for trend includes participants out of range of oestradiol measurements by setting them to the minimum detectable level of 175 pmol/L. Model 2 covariates: age, ethnicity, Townsend deprivation index, alcohol status, daily alcohol intake, days exercised (walked, moderate and vigorous), length of working week, chronotype, job asthma risk and job medical required. Model 3 data are adjusted for Model 2 covariates plus potential mediators sleep duration, smoking status, smoking pack years and BMI.

**Supplementary table 10: Adjusted odds (95% CI) of moderate-severe asthma by current shift work exposure, stratified by sex with day workers as referent. Comparison of models with/without adjusting for testosterone, in participants with measurements for testosterone. (n=216,364)**

| Female/Male |  | Current work schedule | | | Sex-shift work interaction |
| --- | --- | --- | --- | --- | --- |
|  | Day workers | Shift work, but never or rarely night shifts | Irregular shift work including nights | Permanent night shift work |  |
| Total cases (% of total sample size) | 1,466 (1.64%)  1,381 (1.55%) | 179 (1.98%)  152 (1.61%) | 103 (2.12%)  129 (1.40%) | 44 (2.32%)  51 (1.43%) |  |
| Total sample size | 89,575  88,848 | 9,018  9,412 | 4,854  9,190 | 1,900  3,567 |  |
| Model 1: Age-adjusted | Female referent  Male referent | **1.22 (1.04-1.42)**  1.05 (0.89-1.25) | **1.31 (1.07-1.60)**  0.93 (0.78-1.12) | **1.43 (1.05-1.93)**  0.95 (0.72-1.26) | **0.01** |
| Model 1 + testosterone | Female referent  Male referent | **1.21 (1.04-1.42)**  1.05 (0.89-1.25) | **1.30 (1.07-1.60)**  0.93 (0.78-1.12) | **1.43 (1.06-1.94)**  0.95 (0.72-1.26) | **0.01** |
| Model 2: Multivariable-adjusted | Female referent  Male referent | **1.20 (1.02-1.40)**  1.09 (0.92-1.29) | **1.32 (1.08-1.62)**  0.97 (0.81-1.17) | **1.41 (1.04-1.91)**  0.99 (0.74-1.32) | **<0.01** |
| Model 2 + testosterone | Female referent  Male referent | **1.20 (1.02-1.40)**  1.09 (0.92-1.29) | **1.32 (1.08-1.62)**  0.97 (0.80-1.16) | **1.42 (1.04-1.93)**  0.99 (0.75-1.32) | **<0.01** |
| Model 3: Model 2 covariates + potential mediators | Female referent  Male referent | 1.11 (0.95-1.30)  1.06 (0.89-1.26) | 1.22 (0.99-1.50)  0.93 (0.77-1.12) | 1.20 (0.88-1.63)  0.94 (0.70-1.25) | **0.01** |
| Model 3 + testosterone | Female referent  Male referent | 1.10 (0.94-1.29)  1.06 (0.90-1.26) | 1.21 (0.99-1.49)  0.93 (0.78-1.13) | 1.19 (0.87-1.63)  0.94 (0.71-1.25) | **0.01** |

Model 2 covariates: age, ethnicity, Townsend deprivation index, alcohol status, daily alcohol intake, days exercised (walked, moderate and vigorous), length of working week, chronotype, job asthma risk and job medical required. Model 3 data are adjusted for Model 2 covariates plus potential mediators sleep duration, smoking status, smoking pack years and BMI.

**Supplementary table 11: Adjusted odds (95% CI) of moderate-severe asthma by current shift work exposure, stratified by sex with day workers as referent. Comparison of models with/without adjusting for SHBG, in participants with measurements for SHBG. (n=211,866)**

| Female/Male |  | Current work schedule | | | Sex-shift work interaction |
| --- | --- | --- | --- | --- | --- |
|  | Day workers | Shift work, but never or rarely night shifts | Irregular shift work including nights | Permanent night shift work |  |
| Total cases (% of total sample size) | 1,930 (2.08%)  1,279 (1.56%) | 237 (2.51%)  143 (1.65%) | 119 (2.40%)  120 (1.41%) | 62 (3.10%)  48 (1.45%) |  |
| Total sample size | 92,864  82,070 | 9,446  8,691 | 4,956  8,532 | 1,999  3,308 |  |
| Model 1: Age-adjusted | Female referent  Male referent | **1.21 (1.06-1.39)**  1.07 (0.90-1.27) | 1.19 (0.98-1.43)  0.93 (0.77-1.12) | **1.52 (1.18-1.97)**  0.96 (0.72-1.28) | **0.03** |
| Model 1 + SHBG | Female referent  Male referent | **1.20 (1.05-1.38)**  1.07 (0.90-1.27) | 1.17 (0.97-1.42)  0.93 (0.77-1.12) | **1.51 (1.17-1.95)**  0.96 (0.72-1.28) | **0.03** |
| Model 2: Multivariable-adjusted | Female referent  Male referent | **1.18 (1.03-1.36)**  1.11 (0.93-1.32) | 1.19 (0.99-1.44)  0.97 (0.80-1.17) | **1.50 (1.15-1.94)**  1.00 (0.75-1.34) | **0.02** |
| Model 2 + SHBG | Female referent  Male referent | **1.18 (1.02-1.35)**  1.11 (0.93-1.32) | 1.18 (0.98-1.43)  0.97 (0.80-1.17) | **1.49 (1.15-1.93)**  1.00 (0.75-1.34) | **0.02** |
| Model 3: Model 2 covariates + potential mediators | Female referent  Male referent | 1.11 (0.96-1.27)  1.08 (0.91-1.29) | 1.11 (0.92-1.34)  0.93 (0.77-1.14) | **1.31 (1.01-1.70)**  0.95 (0.71-1.28) | **0.03** |
| Model 3 + SHBG | Female referent  Male referent | 1.11 (0.96-1.27)  1.08 (0.91-1.29) | 1.11 (0.92-1.34)  0.93 (0.77-1.14) | **1.31 (1.01-1.70)**  0.95 (0.71-1.28) | **0.03** |

Model 2 covariates: age, ethnicity, Townsend deprivation index, alcohol status, daily alcohol intake, days exercised (walked, moderate and vigorous), length of working week, chronotype, job asthma risk and job medical required. Model 3 data are adjusted for Model 2 covariates plus potential mediators sleep duration, smoking status, smoking pack years and BMI.

**Supplementary table 12: Adjusted odds (95% CI) of moderate-severe asthma by current shift work exposure, stratified by sex with day workers as referent. Comparison of models with/without adjusting for oestradiol, in participants with measurements for oestradiol. (n=49,540)**

| Female/Male |  | Current work schedule | | | Oestradiol-shift work interaction |
| --- | --- | --- | --- | --- | --- |
|  | Day workers | Shift work, but never or rarely night shifts | Irregular shift work including nights | Permanent night shift work |  |
| Total cases (% of total sample size) | 652 (1.92%)  116 (1.57%) | 97 (2.85%)  12 (1.38%) | 54 (2.55%)  9 (1.11%) | 18 (2.37%)  4 (1.27%) |  |
| Total sample size | 33,967  7,372 | 3,404  868 | 2,120  809 | 761  314 |  |
| Model 1: Age-adjusted | Female referent  Male referent | **1.50 (1.21-1.86)**  0.87 (0.48-1.58) | **1.36 (1.03-1.80)**  0.68 (0.34-1.35) | 1.25 (0.78-2.02)  0.78 (0.29-2.13) | 0.16 |
| Model 1 + oestradiol | Female referent  Male referent | **1.50 (1.21-1.86)**  0.87 (0.48-1.58) | **1.36 (1.03-1.80)**  0.68 (0.34-1.36) | 1.25 (0.78-2.02)  0.78 (0.28-2.13) | 0.16 |
| Model 2: Multivariable-adjusted | Female referent  Male referent | **1.47 (1.18-1.82)**  0.91 (0.50-1.67) | **1.37 (1.03-1.82)**  0.70 (0.35-1.40) | 1.25 (0.78-2.02)  0.81 (0.29-2.23) | 0.16 |
| Model 2 + oestradiol | Female referent  Male referent | **1.47 (1.18-1.83)**  0.91 (0.50-1.67) | **1.37 (1.03-1.82)**  0.70 (0.35-1.41) | 1.25 (0.78-2.02)  0.80 (0.29-2.22) | 0.16 |
| Model 3: Model 2 covariates + potential mediators | Female referent  Male referent | **1.35 (1.08-1.69)**  0.94 (0.51-1.72) | 1.29 (0.97-1.71)  0.73 (0.36-1.46) | 1.04 (0.64-1.71)  0.82 (0.30-2.28) | 0.18 |
| Model 3 + oestradiol | Female referent  Male referent | **1.36 (1.09-1.69)**  0.94 (0.51-1.72) | 1.29 (0.97-1.72)  0.73 (0.36-1.47) | 1.04 (0.63-1.70)  0.82 (0.30-2.27) | 0.18 |

Model 2 covariates: age, ethnicity, Townsend deprivation index, alcohol status, daily alcohol intake, days exercised (walked, moderate and vigorous), length of working week, chronotype, job asthma risk and job medical required. Model 3 data are adjusted for Model 2 covariates plus potential mediators sleep duration, smoking status, smoking pack years and BMI.

**Supplementary table 13: Adjusted odds (95% CI) of having moderate-severe asthma by menopause status, defined as self-reported or older than 50, with premenopausal females as referent. (n=221,331)**

|  |  | Menopause Status | | |  |  |
| --- | --- | --- | --- | --- | --- | --- |
|  | Pre-menopausal | Post-menopausal | Unsure – had hysterectomy | Unsure – had hysterectomy (inc. removal of ovaries) | Unsure – other reason | Prefer not to answer |
| Total cases (% of total sample size) | 956 (2.06%) | 4,306 (2.64%) | 121 (3.61%) | 135 (3.56%) | 88 (2.00%) | 2 (1.38%) |
| Total sample size | 46,302 | 163,341 | 3,353 | 3,794 | 4,396 | 145 |
| Model 1: Age-adjusted | Referent | 0.95 (0.85-1.06) | **1.54 (1.26-1.88)** | **1.42 (1.17-1.72)** | 0.92 (0.73-1.16) | 0.76 (0.19-3.06) |
| Model 2: Multivariable-adjusted | Referent | 1.02 (0.91-1.13) | **1.51 (1.24-1.85)** | **1.40 (1.15-1.71)** | 0.91 (0.73-1.15) | 0.54 (0.13-2.24) |
| Model 3: Model 2 covariates + potential mediators | Referent | 0.94 (0.85-1.05) | **1.32 (1.08-1.61)** | 1.21 (0.99-1.47) | 0.84 (0.67 -1.06) | 0.54 (0.13-2.24) |

Model 2 covariates: age, ethnicity, Townsend deprivation index, alcohol status, daily alcohol intake, days exercised (walked, moderate and vigorous), length of working week, chronotype, job asthma risk and job medical required. Model 3 data are adjusted for Model 2 covariates plus potential mediators sleep duration, smoking status, smoking pack years and BMI.

**Supplementary table 14: Adjusted odds (95% CI) of having moderate-severe asthma by menopause status, defined as self-reported or older than 50, with premenopausal females as referent. Participants on HRT or currently taking OCP are excluded. (n=139,822)**

|  |  | Menopause Status | | |  |  |
| --- | --- | --- | --- | --- | --- | --- |
|  | Pre-menopausal | Post-menopausal | Unsure – had hysterectomy | Unsure – had hysterectomy (inc. removal of ovaries) | Unsure – other reason | Prefer not to answer |
| Total cases (% of total sample size) | 849 (2.02%) | 1,929 (2.11%) | 55 (2.62%) | 26 (3.12%) | 69 (1.95%) | 1 (1.41%) |
| Total sample size | 42,023 | 91,250 | 2,098 | 833 | 3,547 | 71 |
| Model 1: Age-adjusted | Referent | 0.88 (0.77-1.00) | 1.23 (0.93-1.64) | 1.36 (0.90-2.05) | 0.92 (0.71-1.18) | 0.78 (0.11-5.60) |
| Model 2: Multivariable-adjusted | Referent | 0.94 (0.82-1.07) | 1.22 (0.92-1.62) | 1.33 (0.88-2.01) | 0.91 (0.70-1.18) | 0.55 (0.08-4.02) |
| Model 3: Model 2 covariates + potential mediators | Referent | 0.90 (0.79-1.03) | 1.07 (0.80-1.42) | 1.07 (0.70-1.65) | 0.83 (0.64 -1.07) | 0.60 (0.08-4.40) |

Model 2 covariates: age, ethnicity, Townsend deprivation index, alcohol status, daily alcohol intake, days exercised (walked, moderate and vigorous), length of working week, chronotype, job asthma risk and job medical required. Model 3 data are adjusted for Model 2 covariates plus potential mediators sleep duration, smoking status, smoking pack years and BMI.

**Supplementary table 15: Adjusted odds (95% CI) of moderate-severe asthma by current shift work exposure in females who have not had a hysterectomy with day workers as referent (n=115,918).**

|  |  | Current work schedule | | |
| --- | --- | --- | --- | --- |
|  | Day workers | Shift work, but never or rarely night shifts | Irregular shift work including nights | Permanent night shift work |
| Total cases (% of total sample size) | 1,961 (1.98%) | 226 (2.31%) | 115 (2.19%) | 60 (2.93%) |
| Total sample size | 98,849 | 9,765 | 5,253 | 2,051 |
| Model 1: Age-adjusted | Referent | **1.17 (1.02-1.35)** | 1.13 (0.93-1.37) | **1.50 (1.16-1.95)** |
| Model 2: Multivariable-adjusted | Referent | 1.14 (0.99-1.32) | 1.14 (0.94-1.39) | **1.48 (1.14-1.93)** |
| Model 3: Model 2 covariates + potential mediators | Referent | 1.07 (0.93-1.24) | 1.06 (0.87-1.28) | 1.30 (0.99-1.69) |

Model 2 covariates: age, ethnicity, Townsend deprivation index, alcohol status, daily alcohol intake, days exercised (walked, moderate and vigorous), length of working week, chronotype, job asthma risk and job medical required. Model 3 data are adjusted for Model 2 covariates plus potential mediators sleep duration, smoking status, smoking pack years and BMI.

**Supplementary table 16: Adjusted odds (95% CI) of having moderate-severe asthma by current shift work exposure with day workers as referent, stratified by menopause status defined as self-reported or older than 50. (n=124,374)**

| Premenopausal  Postmenopausal  Hysterectomy |  | Current work schedule | | | Menopause status-shift work interaction |
| --- | --- | --- | --- | --- | --- |
|  | Day workers | Shift work, but never or rarely night shifts | Irregular shift work including nights | Permanent night shift work |  |
| Total cases (% of total sample size) | 605 (1.87%)  1,300 (2.05%)  291 (2.94%) | 80 (2.53%)  139 (2.21%)  36 (3.00%) | 48 (2.29%)  64 (2.15%)  24 (4.20%) | 21 (2.97%)  37 (2.93%)  12 (4.41%) |  |
| Total sample size | 32,406  63,516  9,910 | 3,164  6,288  1,200 | 2,097  2,981  572 | 706  1,262  272 |  |
| Model 1: Age-adjusted | Premenopausal referent  Postmenopausal referent  Hysterectomy referent | **1.36 (1.08-1.73)**  1.09 (0.91-1.30)  1.02 (0.72-1.45) | 1.24 (0.92-1.66)  1.08 (0.84-1.40)  1.45 (0.95-2.22) | **1.62 (1.04-2.52)**  **1.47 (1.05-2.05)**  1.53 (0.85-2.75) | 0.48 |
| Model 2: Multivariable-adjusted | Premenopausal referent  Postmenopausal referent  Hysterectomy referent | **1.36 (1.07-1.72)**  1.05 (0.88-1.26)  0.98 (0.69-1.40) | 1.28 (0.94-1.73)  1.11 (0.86-1.43)  1.49 (0.97-2.30) | **1.63 (1.04-2.55)**  1.40 (1.00-1.97)  1.43 (0.79-2.61) | 0.47 |
| Model 3: Model 2 covariates + potential mediators | Premenopausal referent  Postmenopausal referent  Hysterectomy referent | 1.27 (1.00-1.62)  0.99 (0.82-1.18)  0.95 (0.67-1.37) | 1.22 (0.90-1.66)  1.02 (0.79-1.32)  1.42 (0.92-2.19) | 1.47 (0.94-2.31)  1.21 (0.86-1.71)  1.33 (0.72-2.45) | 0.54 |

Model 2 covariates: age, ethnicity, Townsend deprivation index, alcohol status, daily alcohol intake, days exercised (walked, moderate and vigorous), length of working week, chronotype, job asthma risk and job medical required. Model 3 data are adjusted for Model 2 covariates plus potential mediators sleep duration, smoking status, smoking pack years and BMI.

**Supplementary table 17: Adjusted odds (95% CI) of having moderate-severe asthma by current shift work exposure with day workers as referent, stratified by menopause status defined as self-reported or older than 50. Participants on HRT or currently taking OCP are excluded (n=86,485)**

| Premenopausal  Postmenopausal  Hysterectomy |  | Current work schedule | | | Menopause status-shift work interaction |
| --- | --- | --- | --- | --- | --- |
|  | Day workers | Shift work, but never or rarely night shifts | Irregular shift work including nights | Permanent night shift work |  |
| Total cases (% of total sample size) | 531 (1.81%)  682 (1.68%)  99 (2.61%) | 72 (2.50%)  69 (1.80%)  11 (2.41%) | 44 (2.26%)  36 (1.91%)  8 (3.45%) | 17 (2.64%)  24 (3.11%)  4 (4.30%) |  |
| Total sample size | 29,282  40,665  3,799 | 2,877  3,834  457 | 1,947  1,885  232 | 643  771  93 |  |
| Model 1: Age-adjusted | Premenopausal referent  Postmenopausal referent  Hysterectomy referent | **1.39 (1.08-1.78)**  1.08 (0.84-1.38)  0.92 (0.49-1.73) | 1.26 (0.92-1.71)  1.16 (0.83-1.63)  1.33 (0.64-2.76) | 1.48 (0.90-2.41)  **1.90 (1.25-2.87)**  1.68 (0.60-4.66) | 0.36 |
| Model 2: Multivariable-adjusted | Premenopausal referent  Postmenopausal referent  Hysterectomy referent | **1.39 (1.08-1.79)**  1.03 (0.79-1.33)  0.90 (0.48-1.71) | 1.31 (0.96-1.80)  1.20 (0.85-1.68)  1.47 (0.69-3.10) | 1.51 (0.92-2.47)  **1.89 (1.24-2.87)**  1.75 (0.62-4.96) | 0.35 |
| Model 3: Model 2 covariates + potential mediators | Premenopausal referent  Postmenopausal referent  Hysterectomy referent | 1.29 (1.00-1.67)  0.97 (0.75-1.26)  0.87 (0.45-1.65) | 1.26 (0.92-1.74)  1.10 (0.78-1.55)  1.35 (0.63-2.86) | 1.35 (0.82-2.22)  **1.60 (1.04-2.46)**  1.68 (0.59-4.82) | 0.42 |

Model 2 covariates: age, ethnicity, Townsend deprivation index, alcohol status, daily alcohol intake, days exercised (walked, moderate and vigorous), length of working week, chronotype, job asthma risk and job medical required. Model 3 data are adjusted for Model 2 covariates plus potential mediators sleep duration, smoking status, smoking pack years and BMI.

**Supplementary table 18: Adjusted odds (95% CI) of having moderate-severe asthma by current shift work exposure with day workers as referent, stratified by sex and menopause status defined as self-reported or older than 50 (n=241,724). Interaction terms consider whether the likelihood of moderate-severe asthma varies differently across shift work frequency categories when comparing males with the three female groups considered separately**

| Male  Female premenopausal/  Female, postmenopausal/  Female, hysterectomy | |  | Current work schedule | | | Sex-shift work interaction  (sub-cohort + males) |
| --- | --- | --- | --- | --- | --- | --- |
|  | Day workers | | Shift work, but never or rarely night shifts | Irregular shift work including nights | Permanent night shift work |  |
| Total cases (% of total sample size) | 1,469 (1.56%)  605 (1.87%)  1,300 (2.05%)  291 (2.94%) | | 161 (1.62%)  80 (2.53%)  139 (2.21%)  36 (3.00%) | 139 (2.38%)  48 (2.29%)  64 (2.15%)  24 (4.20%) | 73 (3.13%)  21 (2.97%)  37 (2.93%)  12 (4.41%) |  |
| Total sample size | 93,937  32,406  63,516  9,910 | | 9,941  3,164  6,288  1,200 | 9,709  2,097  2,981  572 | 3,763  706  1,262  272 |  |
| Model 1: Age-adjusted | Male  referent  Premenopausal referent  Postmenopausal referent  Hysterectomy referent | | 1.05 (0.89-1.23)  **1.36 (1.08-1.73)**  1.09 (0.91-1.30)  1.02 (0.72-1.45) | 0.94 (0.79-1.13)  1.24 (0.92-1.66)  1.08 (0.84-1.40)  1.45 (0.95-2.22) | 0.91 (0.69-1.20)  **1.62 (1.04-2.52)**  **1.47 (1.05-2.05)**  1.53 (0.85-2.75) | **0.03**  0.19  0.14 |
| Model 2: Multivariable-adjusted | Male  referent  Premenopausal referent  Postmenopausal referent  Hysterectomy referent | | 1.08 (0.92-1.28)  **1.36 (1.07-1.72)**  1.05 (0.88-1.26)  0.98 (0.69-1.40) | 0.98 (0.82-1.17)  1.28 (0.94-1.73)  1.11 (0.86-1.43)  1.49 (0.97-2.30) | 0.95 (0.72-1.26)  **1.63 (1.04-2.55)**  1.40 (1.00-1.97)  1.43 (0.79-2.61) | **0.03**  0.16  0.12 |
| Model 3: Model 2 covariates + potential mediators | Male  referent  Premenopausal referent  Postmenopausal referent  Hysterectomy referent | | 1.06 (0.89-1.25)  1.27 (1.00-1.62)  0.99 (0.82-1.18)  0.95 (0.67-1.37) | 0.95 (0.79-1.13)  1.22 (0.90-1.66)  1.02 (0.79-1.32)  1.42 (0.92-2.19) | 0.90 (0.68-1.20)  1.47 (0.94-2.31)  1.21 (0.86-1.71)  1.33 (0.72-2.45) | **0.03**  0.24  0.12 |

Model 2 covariates: age, ethnicity, Townsend deprivation index, alcohol status, daily alcohol intake, days exercised (walked, moderate and vigorous), length of working week, chronotype, job asthma risk and job medical required. Model 3 data are adjusted for Model 2 covariates plus potential mediators sleep duration, smoking status, smoking pack years and BMI.

**Supplementary table 19: Adjusted odds (95% CI) of having moderate-severe asthma by current shift work exposure with day workers as referent, stratified by sex and menopause status defined as self-reported or older than 50. Participants on HRT or currently taking OCP are excluded (n=203,835). Interaction terms consider whether the likelihood of moderate-severe asthma varies differently across shift work frequency categories when comparing males with the three female groups considered separately**

| Male  Female premenopausal/  Female, postmenopausal/  Female, hysterectomy | |  | Current work schedule | | | Sex-shift work interaction (sub-cohort + males) |
| --- | --- | --- | --- | --- | --- | --- |
|  | Day workers | | Shift work, but never or rarely night shifts | Irregular shift work including nights | Permanent night shift work |  |
| Total cases (% of total sample size) | 1,469 (1.56%)  531 (1.81%)  682 (1.68%)  99 (2.61%) | | 161 (1.62%)  72 (2.50%)  69 (1.80%)  11 (2.41%) | 139 (2.38%)  44 (2.26%)  36 (1.91%)  8 (3.45%) | 73 (3.13%)  17 (2.64%)  24 (3.11%)  4 (4.30%) |  |
| Total sample size | 93,937  29,282  40,665  3,799 | | 9,941  2,877  3,834  457 | 9,709  1,947  1,885  232 | 3,763  643  771  93 |  |
| Model 1: Age-adjusted | Male  referent  Premenopausal referent  Postmenopausal referent  Hysterectomy referent | | 1.05 (0.89-1.23)  **1.39 (1.08-1.78)**  1.08 (0.84-1.38)  0.92 (0.49-1.73) | 0.94 (0.79-1.13)  1.26 (0.92-1.71)  1.16 (0.83-1.63)  1.33 (0.64-2.76) | 0.91 (0.69-1.20)  1.48 (0.90-2.41)  **1.90 (1.25-2.87)**  1.68 (0.60-4.66) | 0.06  **0.04**  0.54 |
| Model 2: Multivariable-adjusted | Male  referent  Premenopausal referent  Postmenopausal referent  Hysterectomy referent | | 1.08 (0.92-1.28)  **1.39 (1.08-1.79)**  1.03 (0.79-1.33)  0.90 (0.48-1.71) | 0.98 (0.82-1.17)  1.31 (0.96-1.80)  1.20 (0.85-1.68)  1.47 (0.69-3.10) | 0.95 (0.72-1.26)  1.51 (0.92-2.47)  **1.89 (1.24-2.87)**  1.75 (0.62-4.96) | **0.05**  **0.02**  0.50 |
| Model 3: Model 2 covariates + potential mediators | Male  referent  Premenopausal referent  Postmenopausal referent  Hysterectomy referent | | 1.06 (0.89-1.25)  1.29 (1.00-1.67)  0.97 (0.75-1.26)  0.87 (0.45-1.65) | 0.95 (0.79-1.13)  1.26 (0.92-1.74)  1.10 (0.78-1.55)  1.35 (0.63-2.86) | 0.90 (0.68-1.20)  1.35 (0.82-2.22)  **1.60 (1.04-2.46)**  1.68 (0.59-4.82) | 0.06  **0.05**  0.51 |

Model 2 covariates: age, ethnicity, Townsend deprivation index, alcohol status, daily alcohol intake, days exercised (walked, moderate and vigorous), length of working week, chronotype, job asthma risk and job medical required. Model 3 data are adjusted for Model 2 covariates plus potential mediators sleep duration, smoking status, smoking pack years and BMI.

**Supplementary table 20: Employment characteristics by current shift work exposure (n=285,140)**

|  | Sex | Day workers | Shift work, but never or rarely night shifts | Irregular shift work including nights | Permanent night shift work |
| --- | --- | --- | --- | --- | --- |
| N | F  M | 125,856  109,658 | 12,820  11,606 | 6,793  11,298 | 2,736  4,373 |
| Managers and Senior  Officials (%) | F  M | 13.52  24.96 | 11.72  14.89 | 7.64  10.53 | 3.73  5.97 |
| Professional  Occupations (%) | F  M | 23.64  28.39 | 9.59  11.01 | 8.01  9.50 | 2.74  4.39 |
| Associate Professional and  Technical Occupations (%) | F  M | 17.87  14.46 | 26.95  15.14 | 43.59  20.1 | 38.45  16.33 |
| Administrative and  Secretarial Occupations (%) | F  M | 26.11  6.87 | 12.53  5.18 | 4.78  2.97 | 5.81  3.84 |
| Skilled Trades  Occupations (%) | F  M | 1.57  12.88 | 2.44  15.85 | 1.44  18.68 | 1.02  13.83 |
| Personal Service  Occupations (%) | F  M | 7.91  1.39 | 16.34  5.82 | 25.16  5.85 | 34.25  5.12 |
| Sales and Customer  Service Occupations (%) | F  M | 4.47  1.60 | 11.08  4.58 | 3.61  1.25 | 4.46  2.61 |
| Process, Plant and  Machine Operatives (%) | F  M | 0.90  5.53 | 1.81  15.59 | 1.78  20.76 | 2.27  25.95 |
| Elementary  Occupations (%) | F  M | 4.01  3.91 | 7.53  11.93 | 3.99  10.35 | 7.27  21.95 |

Data are percentages.
